# The effects of quality of evidence communication on perception of public health information about COVID-19: two randomised controlled trials

**DOI:** 10.1101/2021.04.07.21255010

**Authors:** Claudia R. Schneider, Alexandra L. J. Freeman, David Spiegelhalter, Sander van der Linden

## Abstract

**Background:** The quality of evidence about the effectiveness of non-pharmaceutical health interventions is often low, but little is known about the effects of communicating indications of evidence quality to the public.

**Methods:** In two blinded, randomised, controlled, online experiments, US participants (total n=2140) were shown one of several versions of an infographic illustrating the effectiveness of eye protection in reducing COVID-19 transmission. Their trust in the information, understanding, feelings of effectiveness of eye protection, and the likelihood of them adopting it were measured.

**Findings:** Compared to those given no quality cues, participants who were told the quality of the evidence on eye protection was ‘low’, rated the evidence less trustworthy (p=.001), and rated it as subjectively less effective (p=.020). The same effects emerged compared to those who were told the quality of the evidence was ‘high’, and in one of the two studies, those shown ‘low’ quality of evidence said they were less likely to use eye protection (p=.005). Participants who were told the quality of the evidence was ‘high’ showed no significant differences on these measures compared to those given no information about evidence quality.

**Interpretation:** Without quality of evidence cues, participants responded to the evidence about the public health intervention as if it was high quality and this affected their subjective perceptions of its efficacy and trust in the provided information. This raises the ethical dilemma of weighing the importance of transparently stating when the evidence base is actually low quality against evidence that providing such information can decrease trust, perception of intervention efficacy, and likelihood of adopting it.

**Funding:** The Winton Centre for Risk & Evidence Communication, thanks to the David & Claudia Harding Foundation

Research in Context

Evidence before this study
This is the first quantitative, empirical study, to our knowledge, on the effects of communicating the quality of evidence underlying an effectiveness estimate of a public health intervention on a public audience.

Added value of this study
This study provides novel insights into the effects of quality of evidence communication in a public health context. It is thus of high theoretical as well as translational value.

Implications of all the available evidence
Members of the public may assume that information around the effectiveness of a measure such as wearing eye protection to protect against COVID-19 are based on high quality evidence if they are given no cues to suggest otherwise. Yet, when given a statement of the quality of the evidence, this can (appropriately) affect their feelings of the trustworthiness of the information and their subjective judgement of the effectiveness of the measure. This raises the issue of whether there is an ethical imperative to communicate the quality of underlying evidence, particularly when it is low, albeit with the recognition that this may reduce uptake of a public health measure.

## Introduction

Non-pharmaceutical, behavioural interventions such as wearing face coverings, eye protection, and social distancing are a vital part of the mitigations individuals can adopt to protect themselves against infection or transmission of viruses such as SARS-Cov-2. Until an effective vaccine is widely available, such interventions are the only actions that individuals can take themselves [1–3] and they have been described as crucial for successfully managing the pandemic and reducing transmissions if implemented effectively [4–9]. Communication about such interventions, including their effectiveness, to both the public and policy-makers is therefore crucial.

However, despite observational data showing the overall effectiveness of suites of interventions in countries that have mandated various behaviours [4, 5, 7, 9], the evidence around the effectiveness of each potential non-pharmaceutical intervention is still emerging. Attempts at quantification of their effectiveness (e.g. How much does wearing eye protection reduce the chance of infection or transmission of COVID-19?) leads to a number of levels of uncertainty. Any experimental or observational data can give a point estimate (e.g. a percentage point reduction in the chance of infection or transmission) with a confidence interval. Meta-analyses can combine such estimated ranges, but the quantified uncertainty in confidence intervals only reflects a certain amount of the ‘known unknowns’ in an effectiveness estimate. Systematic biases, unexplored variation and a host of other unquantified factors cause deeper uncertainties. These ‘indirect’ uncertainties – not directly about the estimate of effectiveness itself but about the quality of the underlying evidence that the number was derived from – are more difficult to assess and to communicate than a confidence interval [10].

The quality of the evidence base used to produce an estimate of effectiveness plays a crucial role in assessing how reliable and trustworthy the estimate is. If the evidence base is of high quality for instance, the effectiveness information is likely more reliable and less subject to future change compared to when it is derived from low quality evidence. Systems, such as the GRADE working group[11] evidence quality assessments[12], and the Effective Public Health Practice Project Quality Assessment Tool[13] have established ways both of assessing underlying quality of evidence and attempted to produce concise ways to communicate such assessments, via descriptors such as ‘low quality’, ‘moderate quality’ or ‘high quality’ [14, 15].

The limitations in the evidence around COVID-19 mitigation methods means that we are often faced with evidence that – when objectively assessed - is not high quality by any standard: usually rated between ‘very low’ and ‘moderate’ quality by the GRADE. In fact, the same is often true of non-pharmaceutical interventions in other domains, such as physical exercise (e.g. [16]).

This produces a dilemma for public health communicators. Transparent and trustworthy evidence demands clear communication of both effectiveness estimates and the uncertainties around them, including cues of quality of evidence[17]. However, the evidence around the effects of communication of such uncertainty is mixed. Research shows that people prefer transparent communication about scientific uncertainty in general on COVID-19 [18] and communicating quantified uncertainty around an effectiveness estimate (e.g. confidence intervals) often has only very small effects on the public’s overall trust in either the estimate or the source of the message [19, 20]. However, little is known about the effects of communicating the quality of the underlying evidence behind estimates. To address this gap in the literature, in this research, we asked whether communicating the quality of the underlying evidence (in the form of concise labels such as ‘low’ and ‘high’ quality of evidence) affects the public’s trust in the information, their perception of the efficacy of the behavioural intervention being described, and the likelihood of them acting on the conclusions based upon the evidence (e.g. choosing to adopt the intervention).

In June 2020, The Lancet produced an infographic to accompany a meta-analysis of three behavioural interventions to protect against COVID-19 infection or transmission[21]. The infographic used many principles of good evidence communication, with a clear comparison of control and intervention groups with absolute risks in comparable format and an icon array graphic to illustrate the simple percentages. The graphic also included statements about the ‘certainty of evidence’ for each intervention (different from the usual GRADE wording which uses ‘quality of evidence’[14, 15, 22, 23]). The evidence quality ranged from ‘low’ for eye protection and face masks to ‘moderate’ for physical distancing. Whilst all of the other elements of the infographic are based on empirically-tested good practice recommendations for evidence communication (e.g. [24, 25]), the inclusion of cues of certainty or quality of evidence have not been evaluated in this context. We used the Lancet infographic as a real-world setting for tackling the research questions outlined above.

### Overall methods

These experiments were pre-registered (Experiment 1: https://aspredicted.org/blind.php?x=n6pd26; Experiment 2: https://osf.io/ag9th) and given ethical oversight by the Psychology Research Ethics Committee of the University of Cambridge (PRE.2020.086).

US American participants were recruited through the ISO-certified survey panel company Respondi, and were directed to a questionnaire in Qualtrics. Potential participants were asked their age and gender, and a quota system operated to allow only participants who fell into quotas not yet full to continue to the study. The quotas allowed us to match the participants to the US population on age and gender. After giving informed consent, participants answered a series of questions about their COVID-19-related attitudes and beliefs.

#### Randomisation and masking

Participants were then randomised into experimental conditions via the Qualtrics randomisation function. Participants were randomised evenly to each experimental condition and were blinded to the study condition that they were randomized to. All measurements of interest were reported directly by participants rather than being assessed by assessors.

#### Experimental set-up

Once participants entered the experimental groups, they were shown a version of an infographic and asked a series of questions about it. The versions were adapted from the original Lancet infographic described above, which illustrated the evidence for three potential methods of mitigating the transmission of the coronavirus: eye protection, face masks and physical distancing^1^. We chose to study people’s reactions to the presentation of information around eye protection because physical distancing and face masks were both already subject to much public and political discussion in the U.S. at the time of data collection (Sep 24-29, 2020 for Experiment 1; Oct 14-16, 2020 for Experiment 2)[26–29], so we anticipated that the audience may have prior beliefs around both of these measures which may affect their reactions to the experiment. The infographic was experimentally manipulated in Adobe Illustrator to produce different versions for testing.

Our key dependent variables were perceived trustworthiness of the information presented about the effectiveness of eye protection (index of three items, α_Exp1_ = 0.97, α_Exp2_ = 0.96), perceived effectiveness of wearing eye protection, and likelihood of behavioural uptake, i.e. intentions to wear eye protection when in busy public places (each measured on a 7-point Likert scale). See Table 1 for the wording of the dependent measures. We collected further measures for exploratory purposes; the analysis of which are reported in the supplementary materials.

**Table 1:** Overview of dependent measures for Experiments 1 and 2.

| **Concept** | **Questions** | **Answer options** |
| --- | --- | --- |
| Perceived trustworthiness of information | How trustworthy do you think the information you saw on the effectiveness of eye protection is?<br>How accurate do you think the information you saw on the effectiveness of eye protection is?<br>How reliable do you think the information you saw on the effectiveness of eye protection is? | 7 point Likert scale, not trustworthy/accurate/reliable at all – very trustworthy/accurate/reliable |
| Perceived effectiveness of eye protection | How effective do you think eye protection is for reducing the chance of infection or transmission of COVID-19? | 7 point Likert scale, not effective at all – very effective |
| Behavioural uptake intentions | How likely are you to wear eye protection when in busy public places? | 7 point Likert scale, not at all likely – very likely |

The questionnaire for both studies contained an attention check item: “Do you feel that you paid attention, avoided distractions, and took the survey seriously so far?” (answer options: *No, I was distracted; No, I had trouble paying attention; No, I didn’t not take the study seriously so far; No, something else affected my participation negatively; Yes*). Participants who failed the attention check, i.e. who gave an answer other than ‘yes’, were excluded as pre-registered. The attention check measure was administered prior to randomising participants into experimental treatment groups.

All analyses were carried out in R version 3.6

### Experiment 1: Additional Methods

This experiment set out to test whether members of the general public reacted differently to different stated levels of quality/certainty of evidence (high versus low) and to the quality of evidence levels being described as ‘quality of evidence’ (the commonly used GRADE wording) versus ‘certainty of evidence’ (the original Lancet wording), using a 2×2 factorial design (‘low’ versus ‘high’ level of evidence x ‘certainty’ versus ‘quality’ wording) (Figure 1).

**Figure 1:**
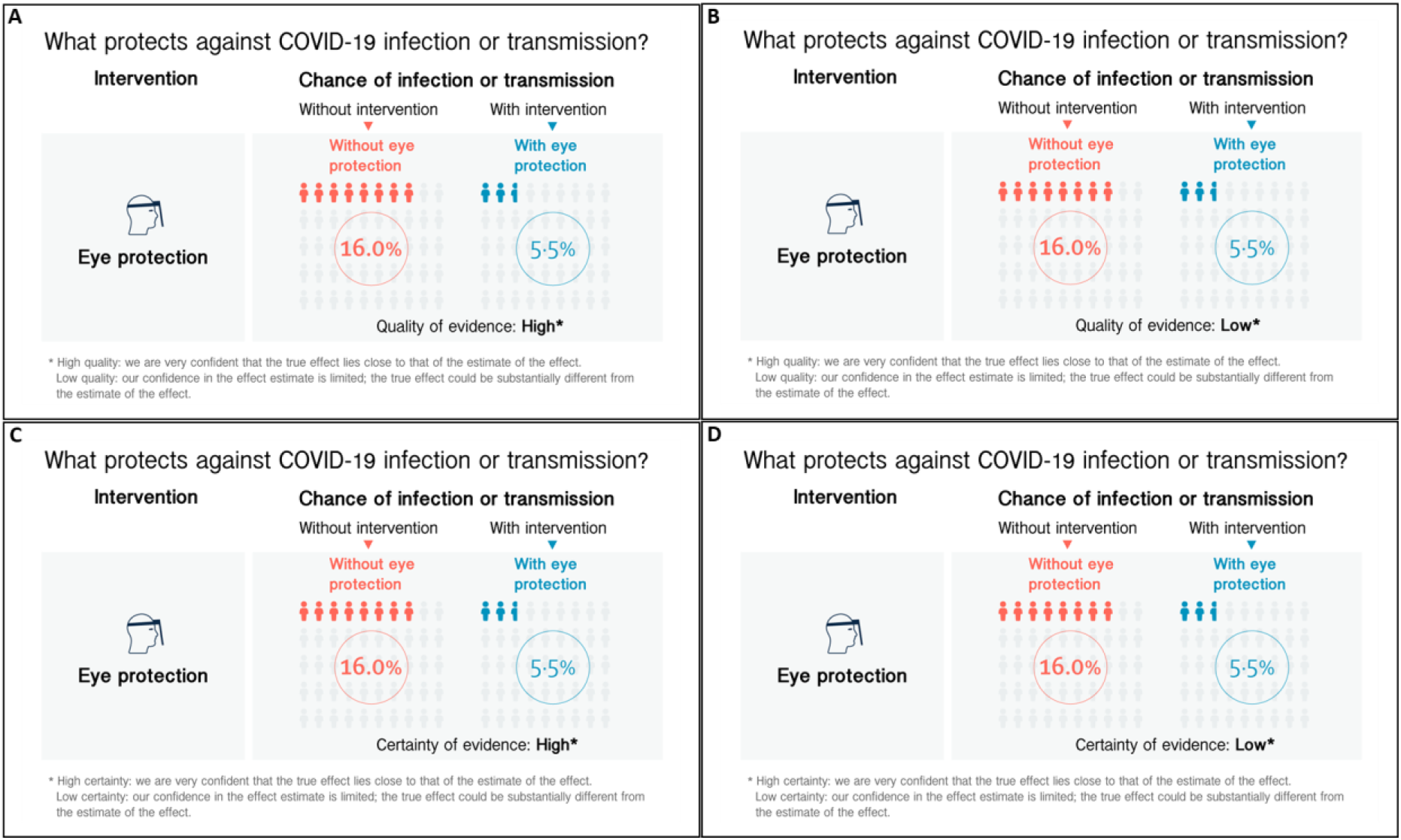
Infographics used in Experiment 1. (A) infographic shown to participants in the High Quality of evidence condition, (B) infographic shown to participants in the Low Quality of Evidence condition, (C) infographic shown to participants in the High Certainty of Evidence condition, (D) infographic shown to participants in the Low Certainty of Evidence condition.

Participants were shown the infographic once, and then asked a series of questions about it on different pages.

We hypothesized (see pre-registration) that people’s trust in the information, their perception of the effectiveness of the intervention, and their likelihood of behavioural uptake would be higher for the group that is shown ‘high’ quality of evidence compared to the group that is shown ‘low’ quality of evidence information.

We pre-registered a sample of 949 participants, providing 95% power at alpha level 0.05 for small effects (f=0.12). This target sample size included a buffer to account for attrition due to failing of the attention check. For data collection, we implemented real-time dynamic sampling which ensured that only those participants who passed the attention check were counted towards the analytic sample quotas. Therefore, the final number of participants for our analytic sample was the full pre-registered sample size.

### Experiment 1: Results

We sampled 949 participants (48.58% male, 51.42% female, *M*_age_ = 45.25, *SD*_age_ = 16.58; see further demographic details as well as number of participants in each experimental condition in the supplementary materials). As pre-registered, we tested for main effects of quality of evidence level and wording for our various outcome measures.

#### Perceived trustworthiness

Two-way Analysis of Variance (ANOVA) revealed a main effect of quality level (‘high’ versus ‘low’) on perceived trustworthiness of the information (F(1,946) = 35.61, p < .001,η_G_^2^ = 0.036). As hypothesized, post-hoc testing using Tukey HSD and Hedge’s correction for effect size adjustment of the contrasts revealed that participants in the low quality of evidence group (*M* = 4.11, 95% CI [3.96,4.26]) indicated significantly lower levels of perceived trustworthiness compared to participants in the high quality of evidence group (*M* = 4.74, 95% CI [4.60,4.88], p < .001, d_adj_ = 0.39, OR = 2.03).

No effect of quality wording (‘certainty of evidence’ versus ‘quality of evidence’) was observed (F(1,946) = 0.01, p = .915).

#### Perceived effectiveness

There was a main effect of quality level on the perceived effectiveness of eye protection

(F(1,946) = 17.4, p < .001, η_G_^2^ = 0.018). Tukey HSD post-hoc tests showed that participants in the low quality of evidence group (*M* = 4.16, 95% CI [4.00,4.33]) indicated significantly lower levels of perceived effectiveness compared to participants in the high quality of evidence group (*M* = 4.64, 95% CI [4.49,4.80], p < .001, d_adj_ = 0.27, OR = 1.63).

There was no main effect of quality wording (F(1,946) = 0.40, p = .528).

#### Behavioural intentions

There was a small significant main effect of quality level for expressed likelihood of behavioural uptake (F(1,946) = 7.80, p = .005, η_G_^2^ = 0.008), such that participants in the low quality of evidence group (*M* = 3.47, 95% CI [3.27,3.66]) indicated significantly lower intentions to wear eye protection compared to participants in the high quality of evidence group (*M* = 3.85, 95% CI [3.66,4.04], p = .005, d_adj_ = 0.18, OR = 1.39).

No main effect of wording emerged (F(1,946) = 0.49, p = .484). Due to the non-normal distribution of the data for the behavioural uptake outcome variable, regular ANOVA was complemented by non-parametric aligned ranks transformation ANOVA which supported the parametric results (main effect of quality of evidence level: F(1,945) = 7.12, p = .008; no effect of wording: F(1,945) = 0.42, p = .518). See Figure 2.

**Figure 2:**
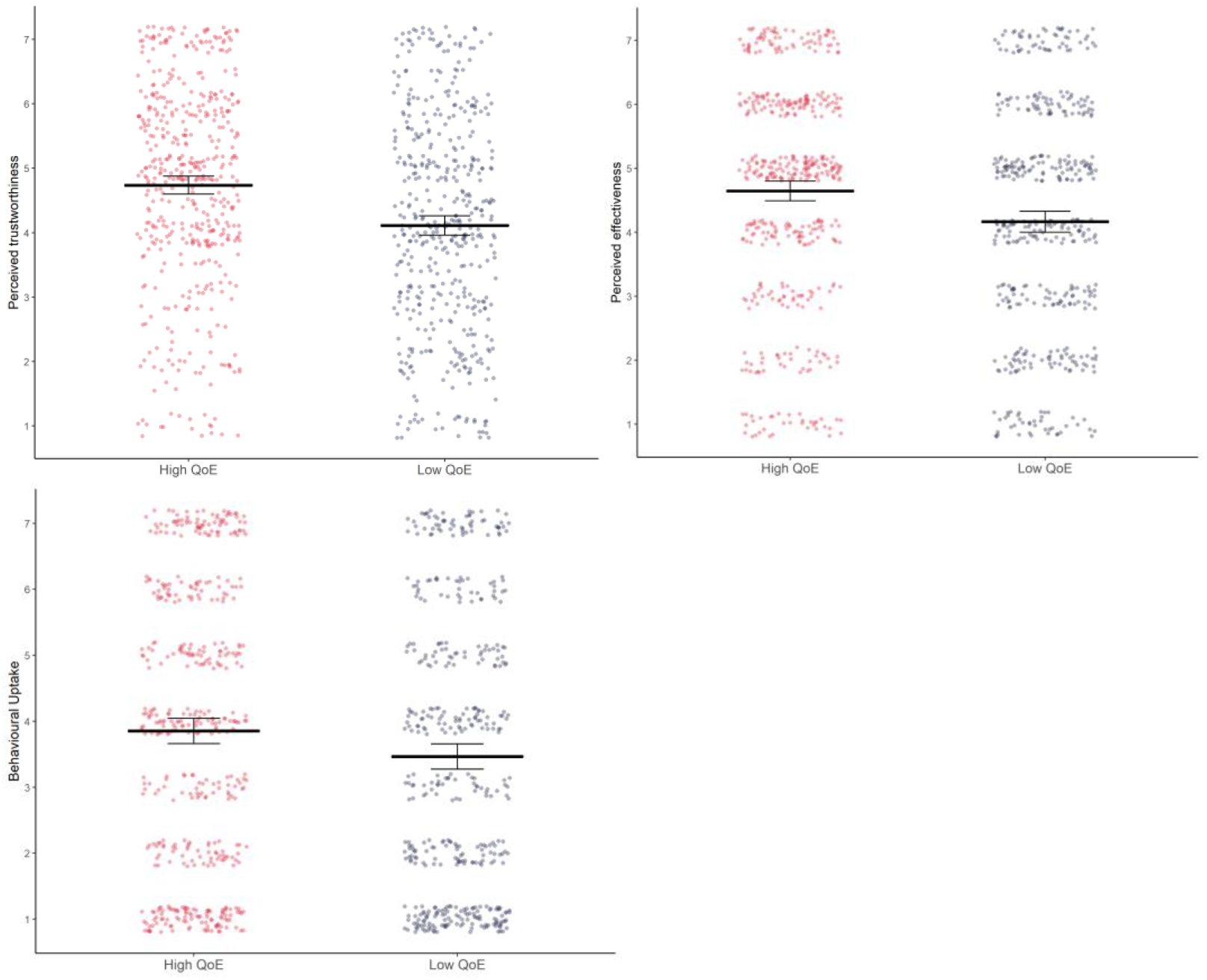
The effects of giving a cue as to the certainty/quality of evidence (QoE) behind an effectiveness estimate on people’s perceptions of the trustworthiness of the information, the perceived effectiveness of the intervention, and the likelihood of them adopting it. Error bars denote 95% confidence intervals. Data is depicted collapsed across wording conditions (quality/certainty).

#### Secondary analysis: understanding

We pre-registered a secondary analysis to explore whether the difference in wording (‘quality’ versus ‘certainty’) affected people’s understanding of the infographic.

Understanding was measured via an index item of reported ease and completeness of comprehension of the effectiveness information in the infographic, as well as self-reported effort invested in understanding the effectiveness information (both measured on a 7 point Likert scale; see supplementary materials for details). An independent samples t-test revealed that ‘quality of evidence’ (*M* = 5.68, 95% CI [5.56,5.80]) was significantly easier to understand for people compared to ‘certainty of evidence’ (*M* = 5.47, 95% CI [5.34,5.60], *t*(939.1) = 2.26, p = .024, d_adj_ = 0.15, OR = 1.31). Since the distribution of the measure was skewed, the parametric analysis was complemented by non-parametric testing for robustness purposes. Mann-Whitney test results were in line with the parametric findings (W = 121759, p = .026). Although descriptively people on average reported lower invested effort for the ‘quality’ wording (*M* = 3.79, 95% CI [3.62,3.97]) compared to the ‘certainty’ wording (*M* = 3.95, 95% CI [3.78,4.13]), this difference was not statistically significant (independent samples t-test: *t*(946) = −1.26, p = .209; Mann-Whitney test: W = 107393,p = .218).

#### Mediation analysis

We had hypothesized that communicating the quality of evidence level would influence people’s perceived trustworthiness of the presented information, which could in turn affect people’s intentions whether to wear eye protection. Specifically, we predicted that providing people with low (versus high) quality of evidence information would decrease people’s trust and hence lead to lower intentions to wear eye protection. To formally test this hypothesis, we pre-registered a mediation analysis of perceived trustworthiness on behavioural uptake intentions.

Mediation analysis was conducted using the mediation package in R [30], with parameter estimates based on 5000 bootstrapped samples for all reported results. Results support our hypothesis: There was a significant direct effect of experimental condition on uptake intentions (b = −0.39, CI [-0.66, −0.12], p = .003) which was no longer significant once the mediator was accounted for (b = 0.09, CI [-0.13, 0.31], p = .418). Importantly, the indirect effect of condition, i.e. low quality of evidence compared to high quality of evidence, on behaviour via perceived trustworthiness was significant (b = −0.48, CI [-0.64, −0.32], p <.001).

#### Additional secondary analyses

We also ran a range of other secondary and exploratory analyses as detailed in the pre-registration. These include the role of reported priors of effectiveness and quality of evidence perception for eye protection, (in-)congruency between priors and presented information, self-reported shifts in trust and behavioural intentions due to the infographic, effects of additional exploratory outcome variables, exploratory interaction analysis between experimental groups, and potential moderators of the observed experimental effects. The results of these additional exploratory analyses can be found in the supplementary materials. As one of these results, we found an interaction between numeracy and quality of evidence level on perceived trustworthiness and effectiveness, hinting at different levels of engagement with, or understanding of, the cues in the infographic between higher numeracy and lower numeracy participants. This finding partially motivated the design of Experiment 2.

### Experiment 1: Discussion

Experiment 1 suggested that there is no difference between participants’ reactions to the words ‘quality’ and ‘certainty’ when used to describe the underlying evidence base behind the use of eye protection in protecting against COVID-19 infection. Although the phrase ‘quality’ could be seen as a more judgemental term (e.g. ‘low quality’ is more pejorative) it appears not to matter whether the term ‘quality’ or ‘certainty’ of evidence is used.

A statement of high quality or certainty of evidence led to the information being trusted more than for a statement of low quality or certainty. Likewise, a statement of high quality or certainty led to people perceiving eye protection as more effective and indicating higher likelihood of wearing eye protection, than for a statement of low quality or certainty.

Additionally, the difference in trust for the high compared to the low quality/certainty condition, appeared to influence the likelihood people said they would wear eye protection.

We therefore proceeded with Experiment 2, leaving aside the ‘certainty’ wording and instead concentrating on investigating the effects of quality of evidence information more deeply. Specifically, we added a crucial condition which had been missing from Experiment 1, i.e. a condition in which people were not given any cues as to the quality of the evidence.

We were also keen to assess the effects of understanding on weighting of information presented in the infographic (i.e. the estimated effectiveness of eye protection and the quality of the evidence the estimate was based on) given the differences in results seen between higher and lower numeracy participants. In reporting health information, it is very common to use numbers (such as percentages) for the effectiveness estimates but simpler, verbal cues for the quality of evidence information. In the Lancet’s infographic, lower numeracy participants’ understanding of the numbers was being supported by graphics in the form of icon arrays. We wondered whether removing this graphical aid, making the information more similar to the standard presentations of experimental results, would make a difference to participants’ reactions to the information given in the infographic.

### Experiment 2: Additional Methods

This experiment had two aims. Firstly, to test how participants assessed evidence quality when there was no statement regarding it, compared to evidence with an overt ‘high’ or ‘low’ quality label. Secondly, to assess whether participants’ reactions to the two quality cues was altered when the icon array was removed, making the efficacy information purely numeric, potentially affecting participants’ understanding.

We therefore used a 3 x 2 factorial design: three conditions of evidence quality cue (‘high’ versus ‘low’ versus no statement) and two conditions of information presentation formats (with and without icon array support to understanding the numbers). See Figure 3.

**Figure 3:**
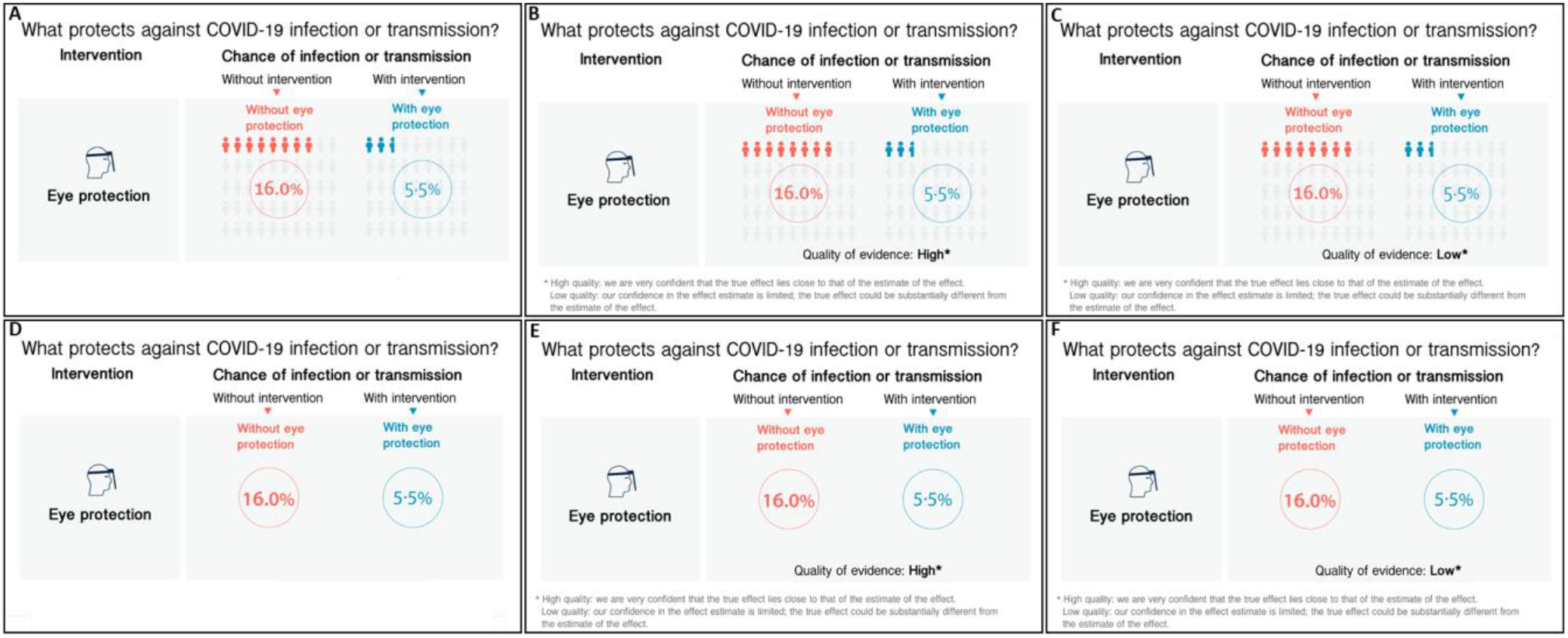
The six infographics used in Experiment 2. (A) infographic shown to participants in the With Graphical Aid No Quality of Evidence group, (B) With Graphical Aid High Quality of Evidence group, (C) With Graphical Aid Low Quality of Evidence group, (D) Without Graphical Aid No Quality of Evidence group, (E) Without Graphical Aid High Quality of Evidence group, (F) Without Graphical Aid Low Quality of Evidence group.

After randomisation, by contrast with Experiment 1, participants were shown the infographic above the questions on each page in this experiment to ensure that all participants had the information in front of them when indicating their responses, reducing potential of a memory recall or ability bias. Key dependent measures were the same as in Experiment 1 (see Table 1).

As for Experiment 1, we hypothesized that people’s trust in the information, their perception of the effectiveness of the intervention, and their likelihood of behavioural uptake would be higher for the group that is shown ‘high’ quality of evidence compared to the group that is shown ‘low’ quality of evidence. Regarding the effects of the condition when people do not receive any quality of evidence information we cautiously hypothesized, based on our experience of ongoing experiments in a different context, that effects of the ‘no quality of evidence’ control group would be closer to the ‘high’ quality of evidence group compared to the ‘low’ quality of evidence group (see pre-registration).

We sampled 1191 participants providing 95% power, at alpha level 0.05 for small effects (f=0.13, based on the results of Experiment 1). We implemented the same real-time sampling procedure checking for attention check fails as described in Experiment 1. The final number of participants for our analytic sample was therefore the full pre-registered sample.

### Experiment 2: Results

We sampled 1191participants (48.53% male, 51.47% female, *M*_age_ = 45.31, *SD*_age_ = 16.43; see further demographic details as well as number of participants in each experimental condition in the supplementary materials). As in Experiment 1, we pre-registered to test for main effects of quality of evidence level and format for our various outcome measures.

#### Perceived trustworthiness

Two-way analysis of variance using Tukey HSD and Hedge’s correction for effect size adjustment of the contrasts revealed a main effect of quality of evidence level on perceived trustworthiness of the information (F(2,1187) = 15.05, p < .001, η_G_^2^ = 0.025), such that participants in the low quality of evidence group (*M* = 4.07, 95% CI [3.91,4.24]) indicated significantly lower levels of perceived trustworthiness compared to participants in the group that did not present quality of evidence information at all (*M* = 4.48, 95% CI [4.32,4.63], p = .001, d_adj_ = 0.25, OR = 1.57), as well as compared to those in the high quality of evidence group (*M* = 4.68, 95% CI [4.53,4.84], p < .001, d_adj_ = 0.38, OR = 1.99). Participants in the high quality of evidence group did not significantly differ in their trust from those in the group that did not receive quality of evidence information (p = .169).

No main effect of format emerged (F(1,1187) = 0.53, p = .465).

#### Perceived effectiveness

A main effect of quality of evidence level on perceived effectiveness emerged (F(2,1187) = 9.98, p < .001, ηG2 = 0.017), such that participants in the low quality of evidence group (*M* = 4.11, 95% CI [3.94, 4.29]) indicated significantly lower levels of perceived effectiveness compared to those participants who were not presented with quality of evidence information (*M* = 4.45, 95% CI [4.27, 4.62], p = .018, d_adj_ = 0.19, OR = 1.41), as well as compared to participants in the high quality of evidence group (*M* = 4.65, 95% CI [4.49, 4.81], p < .001, d_adj_ = 0.31, OR = 1.75). The high quality of evidence group was not significantly different from the no quality of evidence information group (p = .203). See Figure 4.

**Figure 4:**
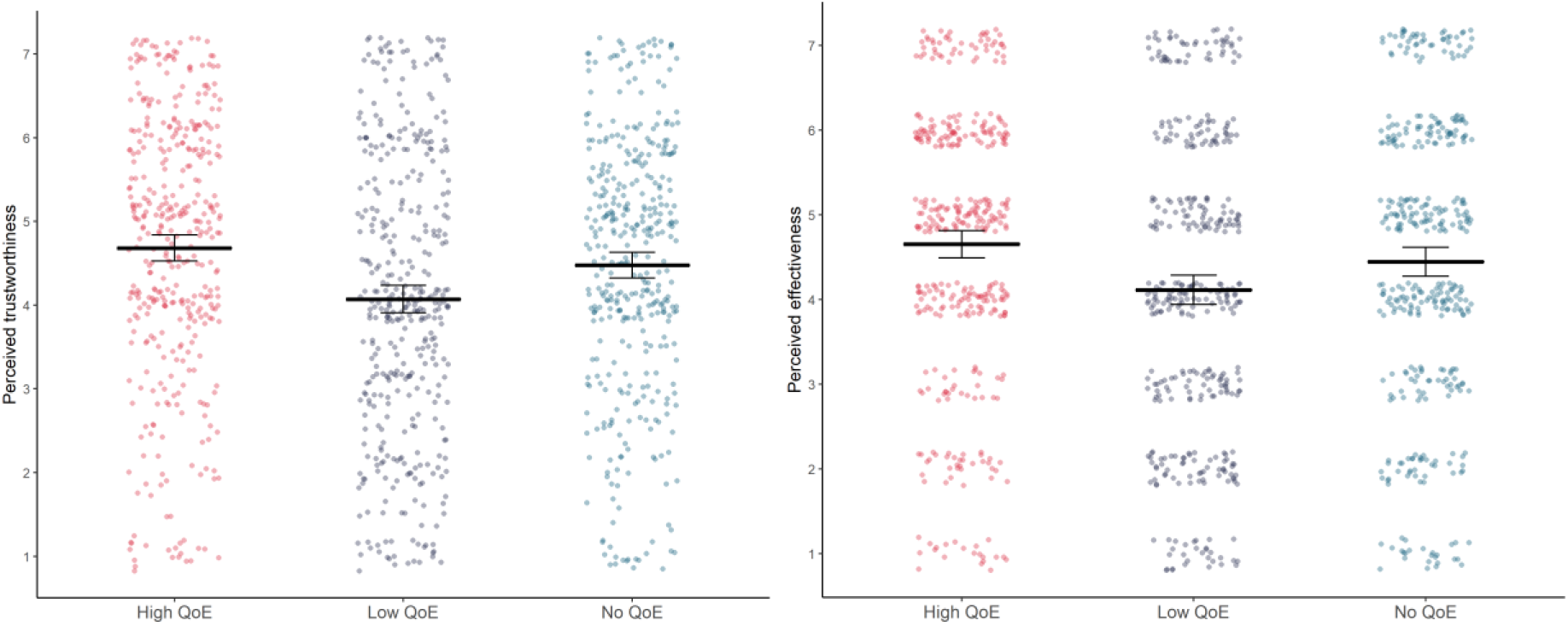
The effects of giving a cue as to the quality of evidence (QoE) behind an effectiveness estimate on people’s perceptions of the trustworthiness of the information and the perceived effectiveness of the intervention. Error bars denote 95% confidence intervals. Data is depicted collapsed across formatting conditions (percentages/percentages and icon arrays).

No main effect of format was observed (F(1,1187) = 2.68, p = .102).

#### Behavioural intentions

No significant effects emerged for the behavioural uptake measure, either for quality of evidence level (F(2,1187) = 1.28, p = .279) or for format (F(1,1187) = 0.46, p = .496). Because of the non-normal distribution of the data for the behavioural uptake measure, regular ANOVA was complemented by non-parametric aligned ranks transformation ANOVA which did not reveal any significant effects either (quality of evidence level: F(2,1185) = 1.17, p = .311; format: F(1,1185) = 0.01, p = .905).

#### Understanding

We had hoped to explore the potential influence of people’s understanding of the information given in the infographic, especially its role in shaping the effects of the various levels of quality of evidence information on trust, perceived effectiveness and behaviour. However, a check to see whether our experimental manipulation of the infographic (by removal of the icon array) had made a significant difference to participants’ self-reported understanding of the information (index item of reported ease and completeness of comprehension of the effectiveness information in the infographic) revealed that it had not (*t*(1188.9) = 0.04, p = .970; Wilcoxon rank sum test, W = 177244, p = .992).

For completeness, we still ran our pre-registered interaction analyses between quality of evidence level and format, as well as self-reported understanding. We investigated people’s self-reported ease and completeness of understanding of the effectiveness information in the infographic, their objective understanding of the numeric effectiveness information, as well as the amount of effort they reported to have invested in understanding the information on the effectiveness of eye protection in the infographic. We did not find any significant interactions for any of these potential moderators on any of our outcome measures. Detailed results and those of further exploratory analyses are reported in the supplementary materials.

### Experiment 2: Discussion

Experiment 2 replicated the main effects on two of our three dependent variables: there was a significant effect of giving quality of evidence information on both the perceived trustworthiness of the information and the perceived effectiveness of the intervention. No significant differences emerged for the behavioural uptake measure.

Experiment 2 furthermore extended the findings from Experiment 1 in important ways. In addition to replicating effects of providing high versus low quality of evidence information, we tested how people would react if not given any quality of evidence information at all. As hypothesized, we found that the effects of not giving people indications of the quality level of the evidence were similar to those seen in the high quality of evidence group, and significantly different from those of the low quality of evidence group.

This suggests that in the absence of external cues of the quality of the evidence, people responded to the information they were provided with as if it was high quality, and that only stating overtly that evidence is ‘low quality’ could significantly change people’s perceptions. This might be because people implicitly assume a relatively high level of quality of evidence they are presented with in this kind of scenario (i.e., in the absence of external cues of its quality), or because their response to anything other than explicit low quality is not significantly different from their response to high quality. The latter could be an example of loss aversion. Psychological theory has shown that people are more sensitive to losses compared to gains, which may be why they react more strongly to low quality of evidence information compared to high quality of evidence information which pulls effects downwards for the ‘low’ group [31–33].

When looking at the effects of degree of understanding on weighting of cues, unfortunately our manipulation of the format did not make a significant difference to the understandability of the numerical effectiveness information, which implies that the icon arrays were not making as much difference as we’d hoped in supporting the comprehension of the numbers presented.

## Conclusions

Across two large, randomised experiments we show that information about the quality of underlying evidence changes public perceptions of estimates of the effectiveness of public health measures.

In Experiment 1 we show that a statement of high quality or certainty of evidence led to the information being trusted more than for a statement of low quality or certainty. In the same way it also affected how effective people judged eye protection to be in reducing the chance of COVID-19 infection, and the likelihood to which people indicated they would wear eye protection. Moreover, we show that effects on trust mediate the relationship between quality of evidence information and downstream behaviour. We find that providing people with low (versus high) quality of evidence information decreased people’s trust and in turn lowered their intentions to wear eye protection.

Looking at different ways of referring to quality of evidence information (Experiment 1), we find no difference between participants’ reactions to the words ‘quality’ and ‘certainty’ on measures of trust, perceived effectiveness or behavioural uptake intentions, although the two words are qualitatively different. As people showed differential responses to the level of quality of evidence, it may be that they pay more attention to, or weight more, the qualifier (e.g. ‘low’ or ‘high’) than the terminology of the measure. We do however find effects on understanding, such that participants rated the term ‘quality of evidence’ to be easier to understand compared to ‘certainty of evidence’ and suggest that communicators use the term ‘quality’.

Understanding people’s reactions to public health communications gives important insights on factors affecting compliance and ultimately the success of non-pharmaceutical interventions. Although several studies have assessed the effects of non-pharmaceutical interventions in the context of COVID-19 in various countries [4–9], these studies have largely relied on modelling approaches using observational data, such as information on lockdown measures and other imposed restrictions and measures of COVID-19 prevalence (e.g. reproduction rates) [4, 5, 7, 9]. As we show through experimental randomised controlled trials, people’s reactions to public health communication critically depend on their perceptions about whether they are being presented with high or low quality information, and this affects how likely they say they are to take action based on the communication.

By contrast with our findings here about ‘indirect’ uncertainties around the quality of the evidence underlying numerical estimates, experiments on the communication of ‘direct’ uncertainty – uncertainty around the actual numerical estimate itself – appears to have much less effect on trust, and full disclosure is preferred by the public [18, 19].

To our knowledge, this is the first published evidence on the effects of communicating ratings of the quality of evidence around health-related findings and as a result, an important ethical issue emerges from our findings. In the absence of cues around quality, our evidence here suggests that people treat the information they are provided with as if it is based on high quality evidence, and this affects their reactions to it. If the facts being communicated to the public are actually known to be based on low quality evidence, lack of disclosure of this has implications: it could be seen to be misleading.

In the case of individual medical decisions, where information is being given purely as a matter of informed consent or shared decision-making, the ethical (and sometimes legal) implications are clear: disclosure of the quality of the underlying evidence base is vital. However, in the realm of public health, where the mandate may be more to persuade than inform, it may be tempting for communicators to not disclose the low quality of evidence levels in order to promote compliance. However, that is a decision that has to be made in the knowledge that that lack of disclosure is likely to affect people’s reactions to the information.

This study is limited in that it tested only an online population in the US (albeit quota sampled), and only one health intervention. Further research could broaden this population and context. It would also be useful to examine the effects of providing greater nuance and detail on the quality rating, and effects on different subgroups of the population, such as low and high numeracy individuals.

## Data Availability

All data collected for the reported studies (de-identified participant data), along with the questionnaires used, is publicly available at https://osf.io/z6ps9/

## Authors’ contributions

Claudia R. Schneider: Conceptualization, Methodology, Data curation, Formal analysis, Investigation, Project administration, Validation, Visualization, Writing – original draft, Writing – review & editing

Alexandra L. J. Freeman: Conceptualization, Methodology, Validation, Writing – original draft, Writing – review & editing

David Spiegelhalter: Conceptualization, Methodology, Writing – review & editing

Sander van der Linden: Conceptualization, Methodology, Validation, Writing – review & editing

## Footnotes

1 https://www.eurekalert.org/multimedia/pub/233365.php; https://www.eurekalert.org/pub_releases/2020-06/tl-pss060120.php; https://www.thelancet.com/journals/lancet/article/PIIS0140-6736(20)31142-9/fulltext

