## Supplementary material for "The effects of quality of evidence communication on perception of public health information about COVID-19: two randomised controlled trials"

April 2021

Corresponding author: Claudia R. Schneider, Winton Centre for Risk and Evidence Communication, University of Cambridge, Wilberforce Road, Cambridge, United Kingdom.


### Experiment 1

#### Demographic composition of the study sample

| Variable | **Study 1** ($N$ = 949) |
|---|---|
| Gender, % | |
| Females | 51.42 |
| Males | 48.58 |
| Age, $M$ ($SD$) | 45.25 (16.58) |
| Education, % | |
| Did not complete high school | 1.69 |
| High school degree or equivalent | 35.83 |
| Associate's degree | 14.44 |
| Bachelor's degree | 32.03 |
| Graduate or Professional degree | 16.02 |
| Political views, $M$ ($SD$) | 4.05 (1.67) |

*Political views on spectrum from very left wing (or liberal) to very right wing (or conservative) on 7-point scale.

#### Infographic used and sample size per experimental condition

High quality of evidence (n = 240):

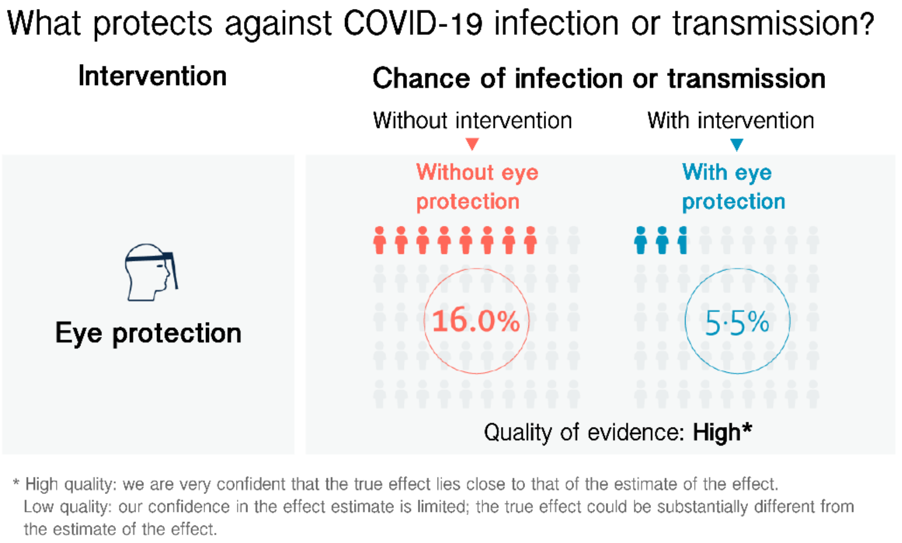

Low quality of evidence (n = 243):

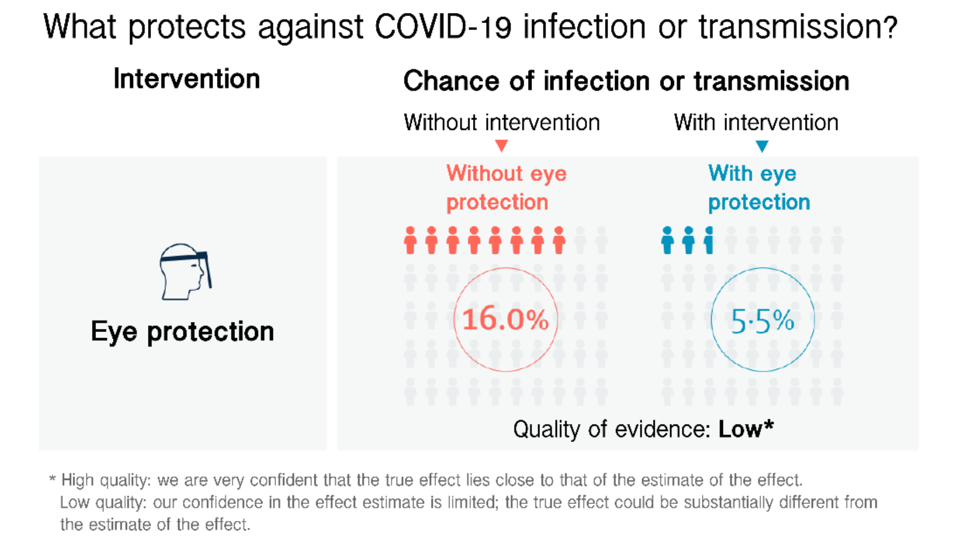

High certainty of evidence (n = 243):

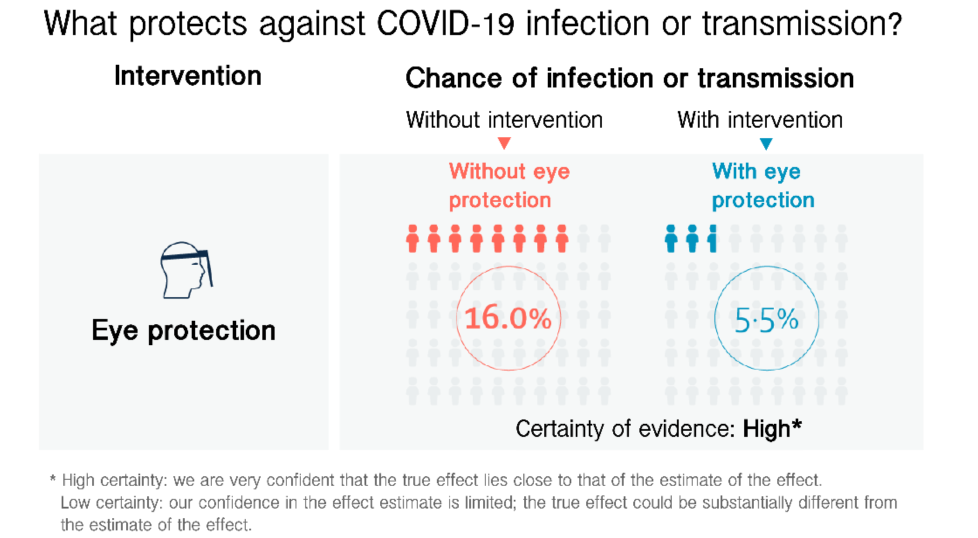

Low certainty of evidence (n = 223):

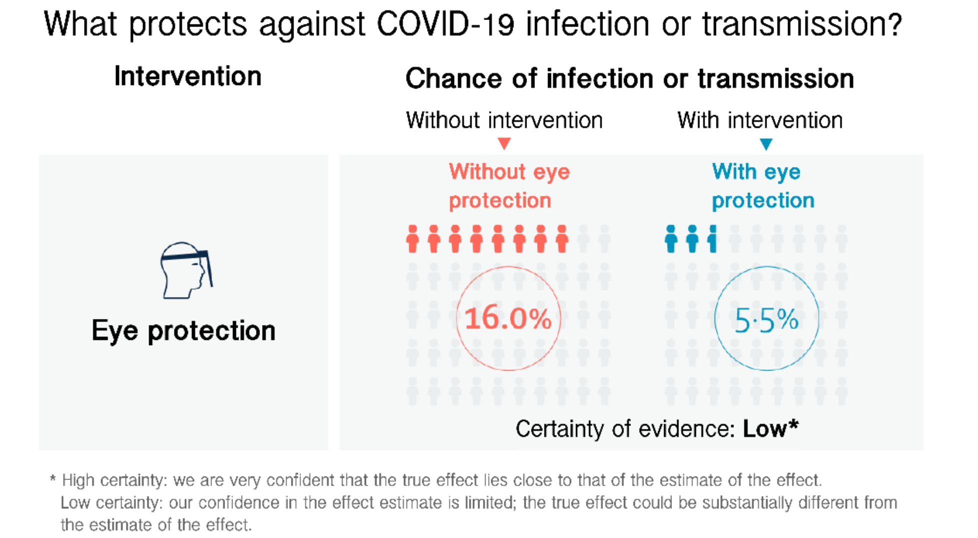

#### Survey measures

**Dependent measures:**
Perceived trustworthiness index items (α = 0.97):
How accurate do you think the information you saw on the effectiveness of eye protection is? Not accurate at all (1) - Very accurate (7)
How reliable do you think the information you saw on the effectiveness of eye protection is? Not reliable at all (1) - Very reliable (7)
How trustworthy do you think the information you saw on the effectiveness of eye protection is? Not trustworthy at all (1) - Very trustworthy (7)

Perceived effectiveness:
How effective do you think eye protection is for reducing the chance of infection or transmission of COVID-19? Not effective at all (1) - Very effective (7)

Behavioural uptake:
How likely are you to wear eye protection when in busy public places? Not at all likely (1) - Very likely (7)

**Understanding and effort questions:**
Index of ease and completeness of understanding (r = 0.74, rho = 0.76):
How easy or difficult did you find the information on the effectiveness of eye protection to understand? Very difficult (1) - Very easy (7)
How completely do you feel you understood the information on the effectiveness of eye protection? Not at all (1) - Completely (7)

Effort invested:
How much effort do you feel you had to put into understanding the information on the effectiveness of eye protection? None (1) - A lot (7)

#### Model diagnostics

Quality of evidence level and wording on perceived trustworthiness:

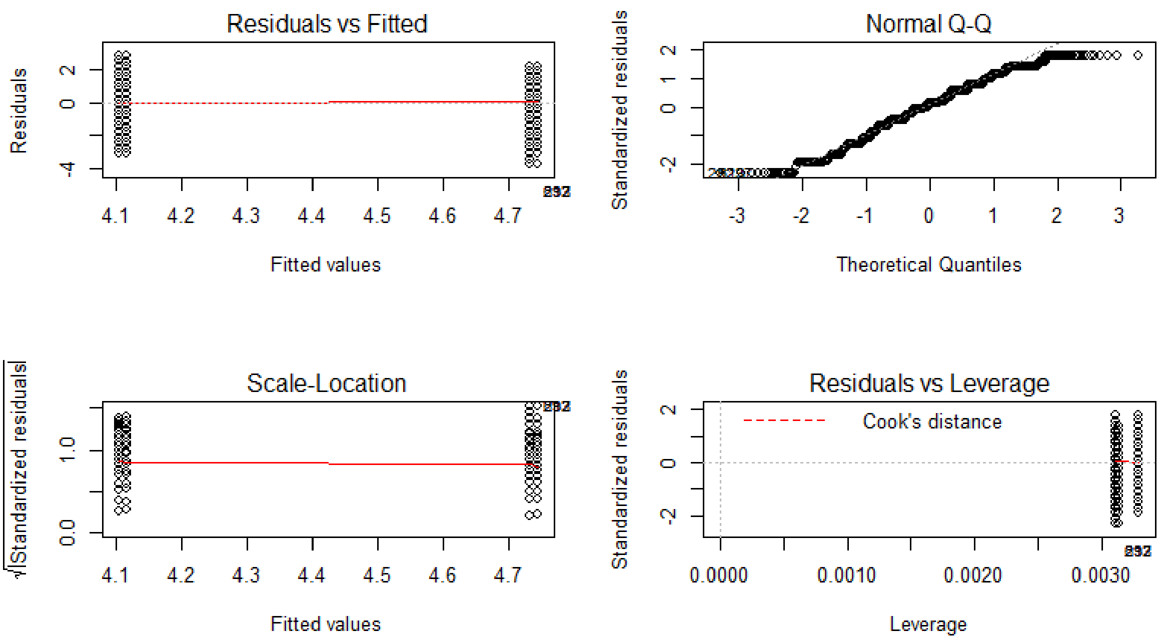

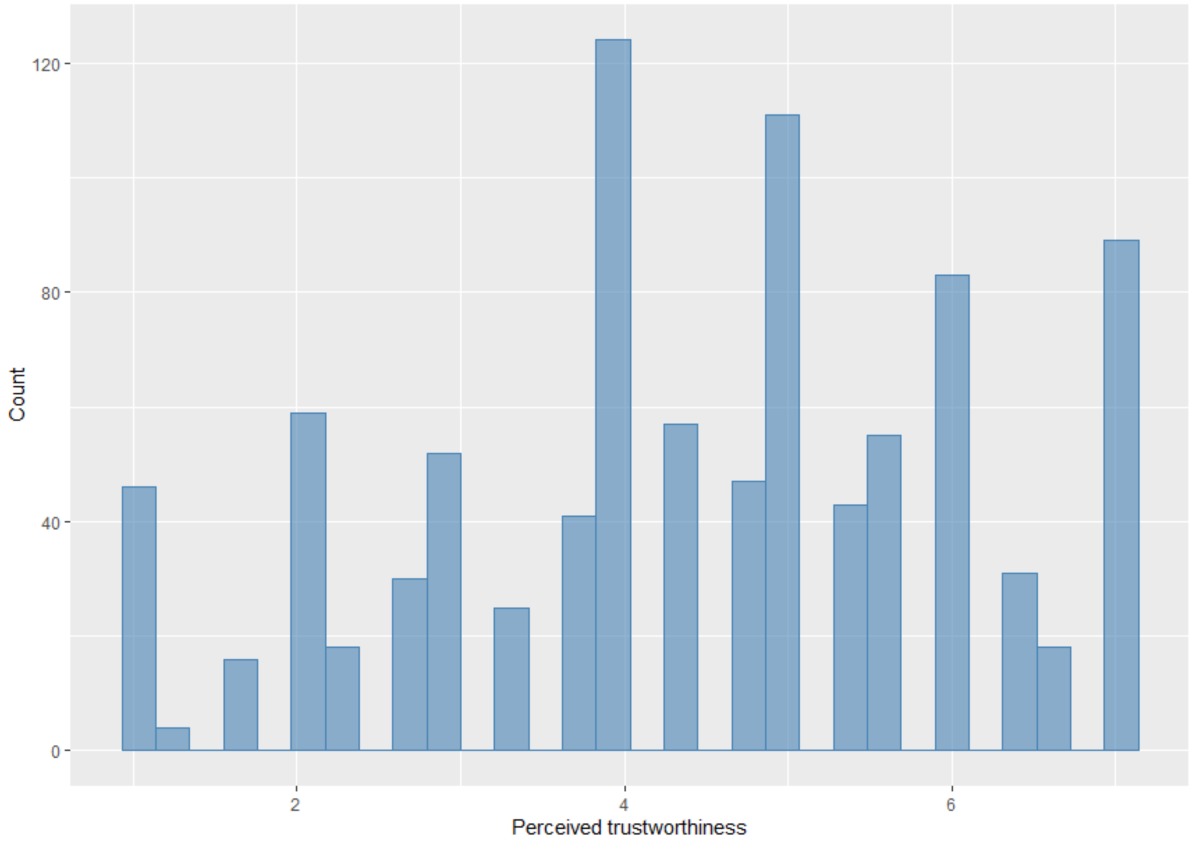

Quality of evidence level and wording on perceived effectiveness:

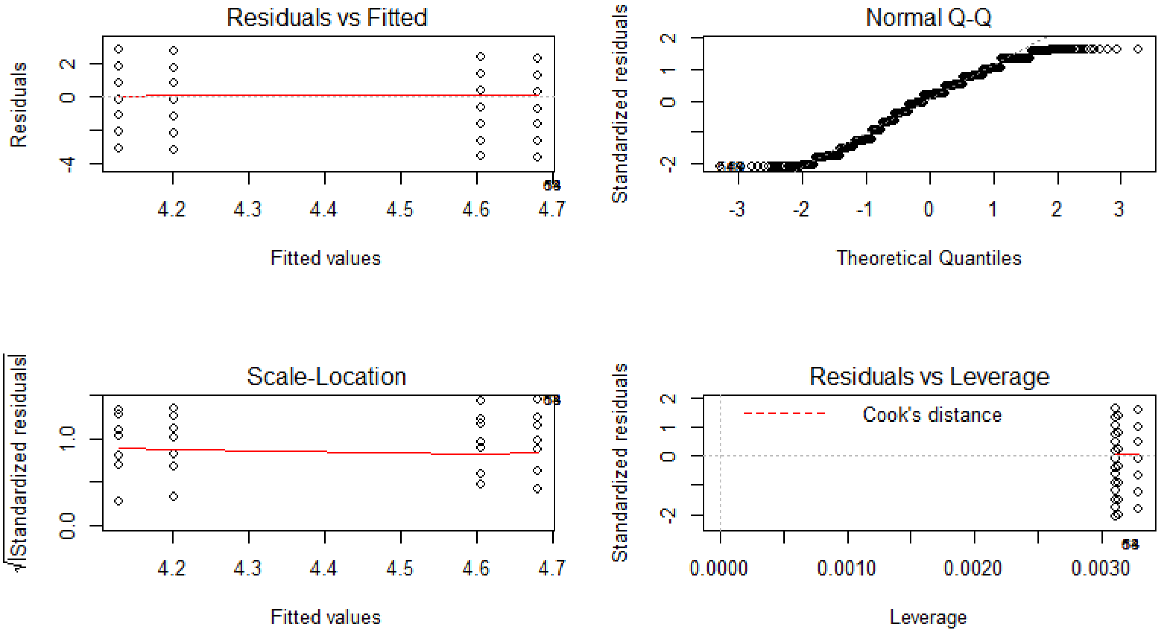

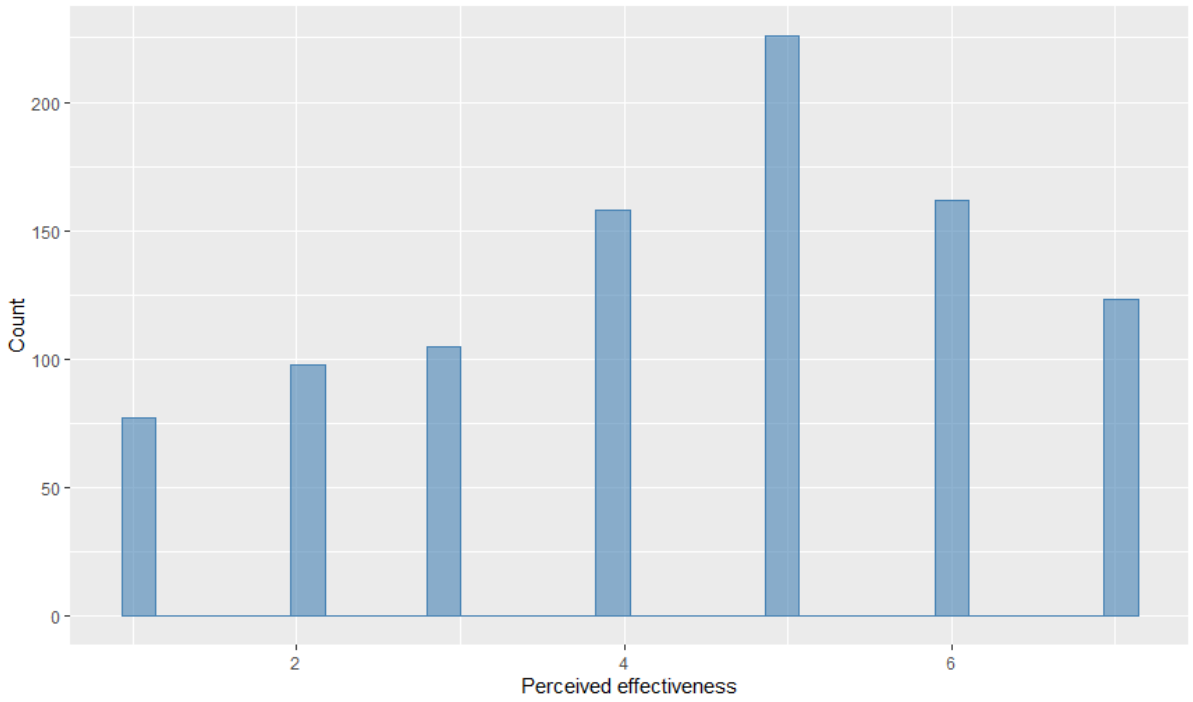

Quality of evidence level and wording on behavioural uptake intentions:

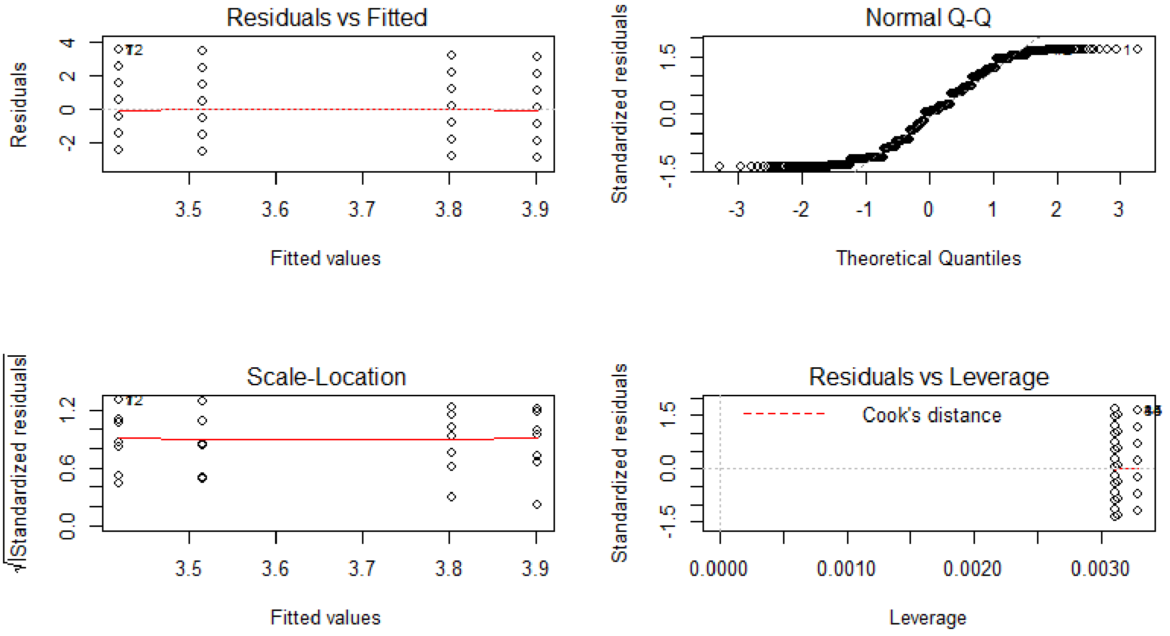

Note: Non-parametric aligned rank transformed ANOVA was conducted, due to skew in the outcome measure.

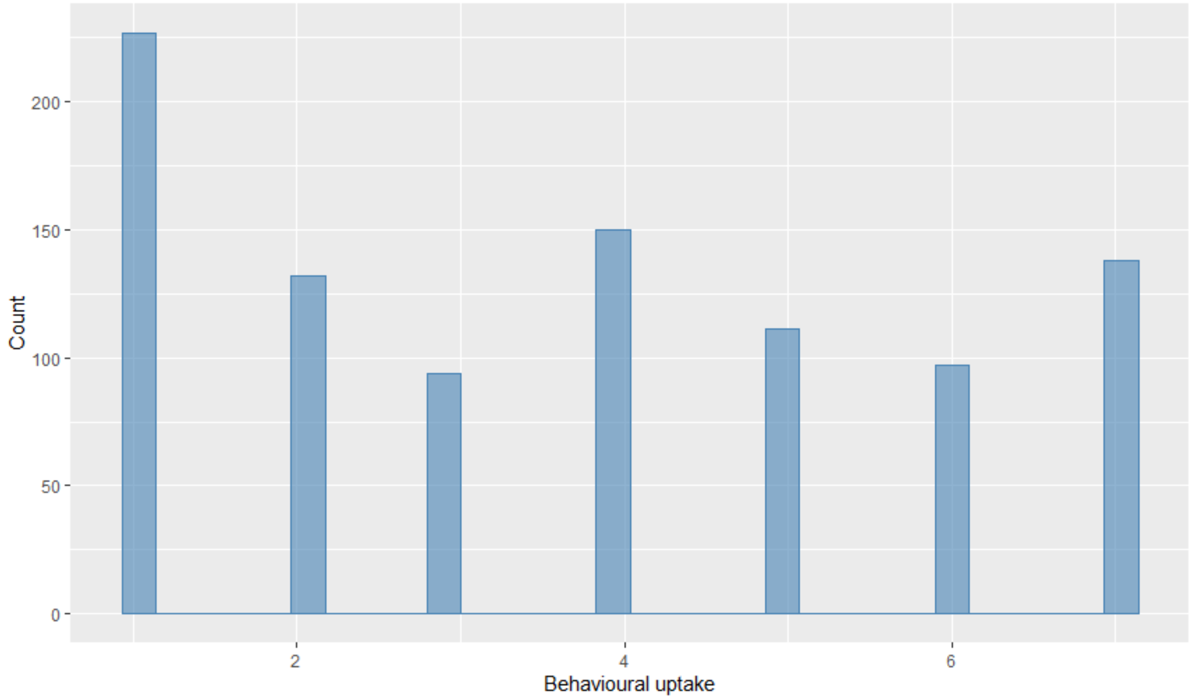

Quality of evidence wording on ease and completeness of understanding:

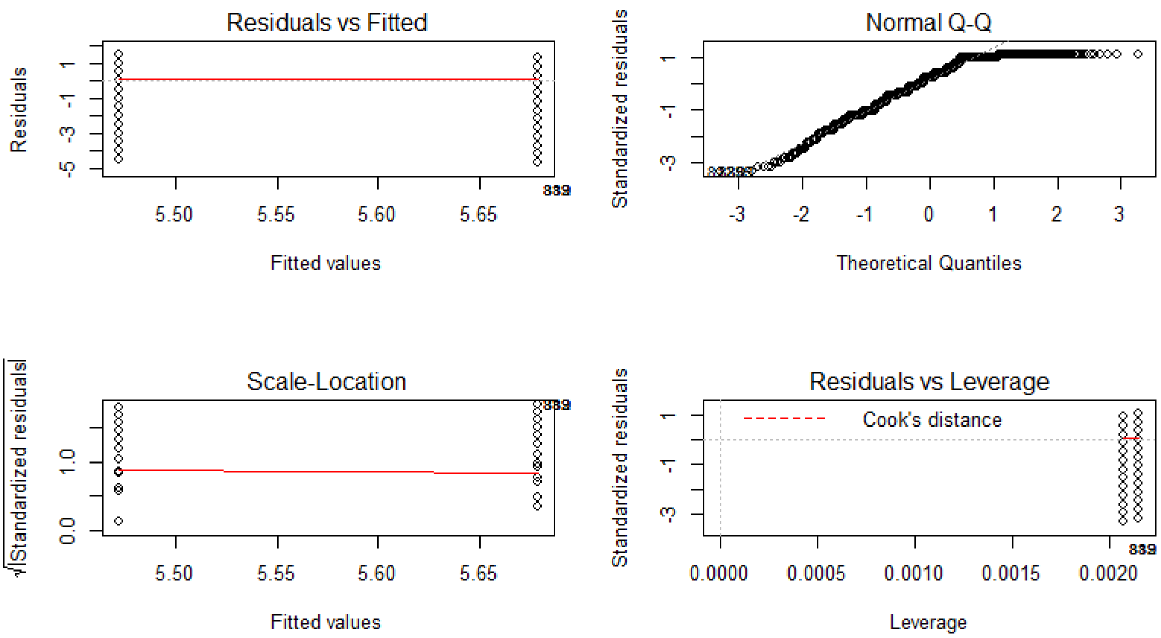

Note: Due to skew in the outcome variable, non-parametric analysis (in addition to parametric analysis) was performed for robustness purposes.

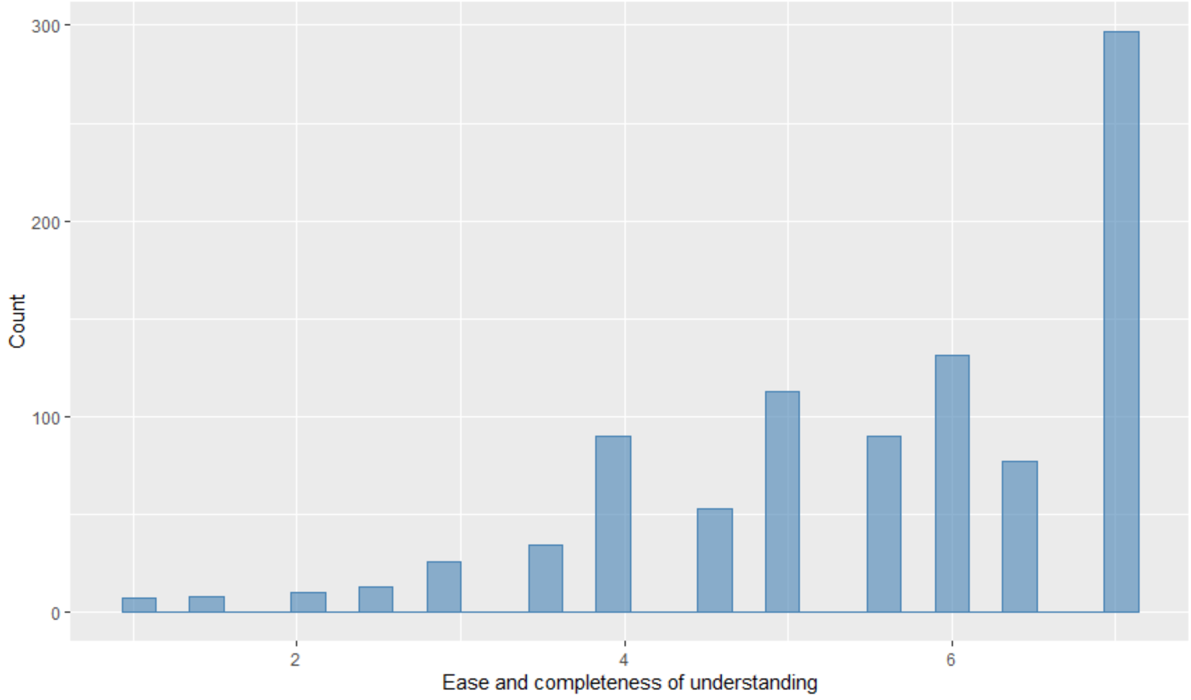

Quality of evidence wording on effort invested for understanding:

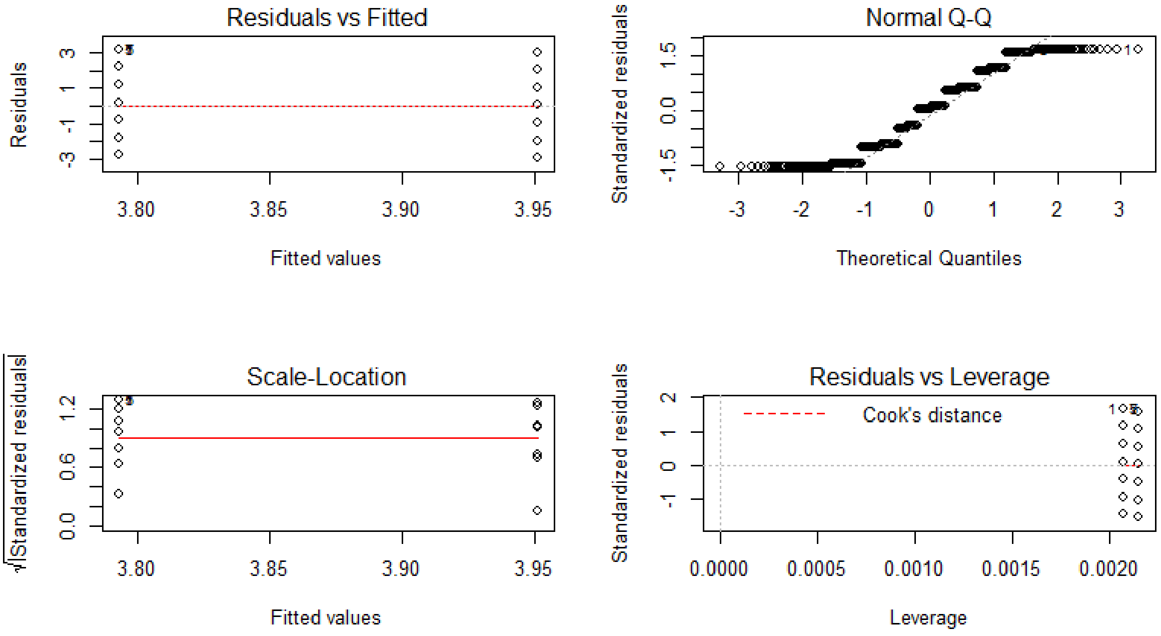

Note: Due to the non-normality of the outcome variable, non-parametric analysis (in addition to parametric analysis) was performed for robustness purposes.

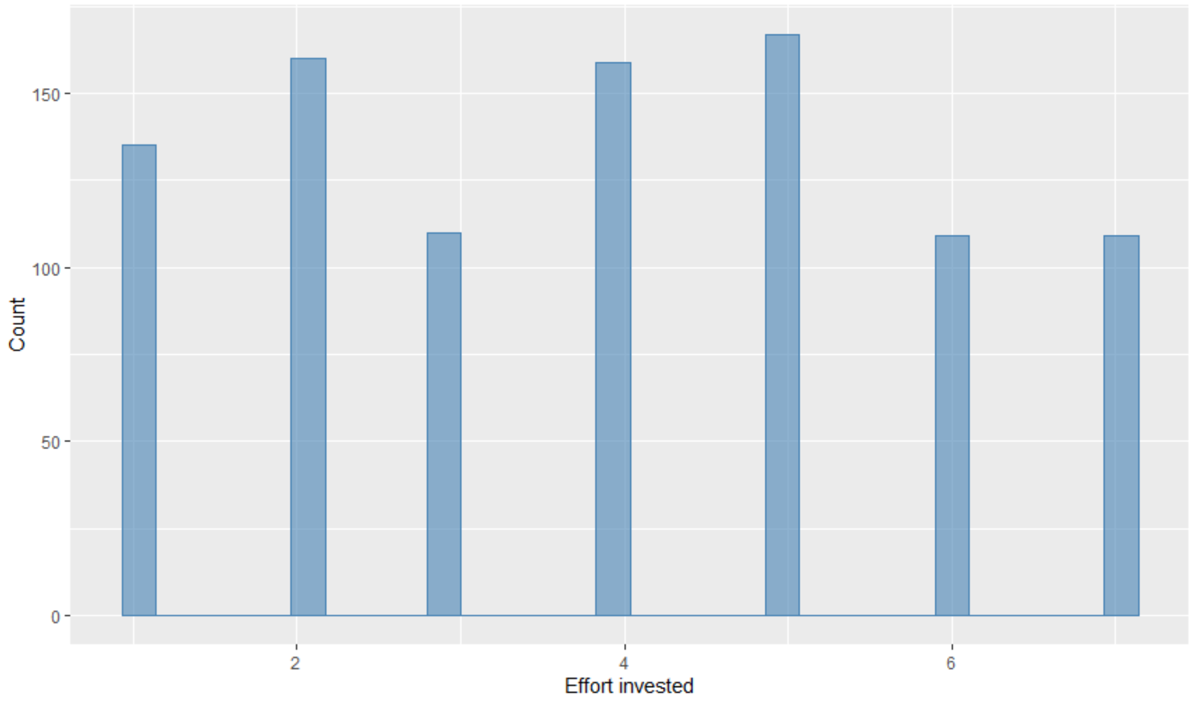

### Experiment 2

#### Demographic composition of the study sample

| Variable | **Study 2** (*N* = 1,191) |
|---|---|
| Gender, % | |
| Females | 51.47 |
| Males | 48.53 |
| Age, *M* (*SD*) | 45.31 (16.43) |
| Education, % | |
| Did not complete high school | 2.18 |
| High school degree or equivalent | 38.20 |
| Associate's degree | 15.28 |
| Bachelor's degree | 29.39 |
| Graduate or Professional degree | 14.95 |
| Political views, *M* (*SD*) | 3.99 (1.62) |

*Political views on spectrum from very left wing (or liberal) to very right wing (or conservative) on 7-point scale.

#### Infographic used and sample size per experimental condition

High quality of evidence – percentages only (n = 196):

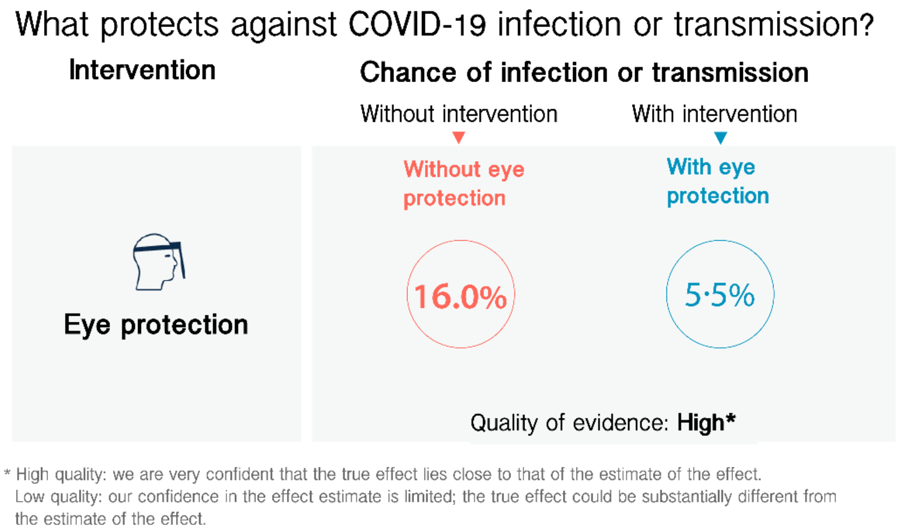

High quality of evidence – percentages and icon arrays (n = 202):

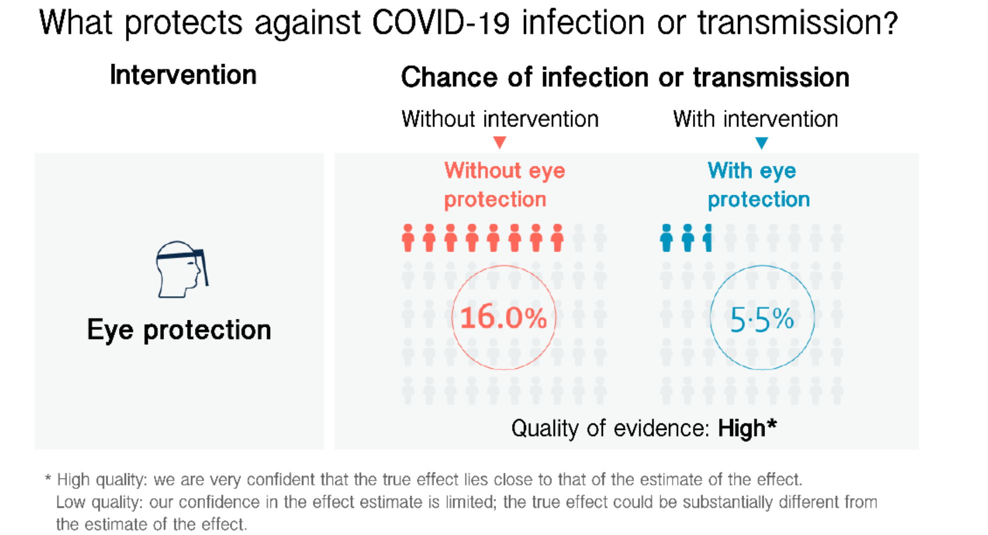

Low quality of evidence – percentages only (n = 204):

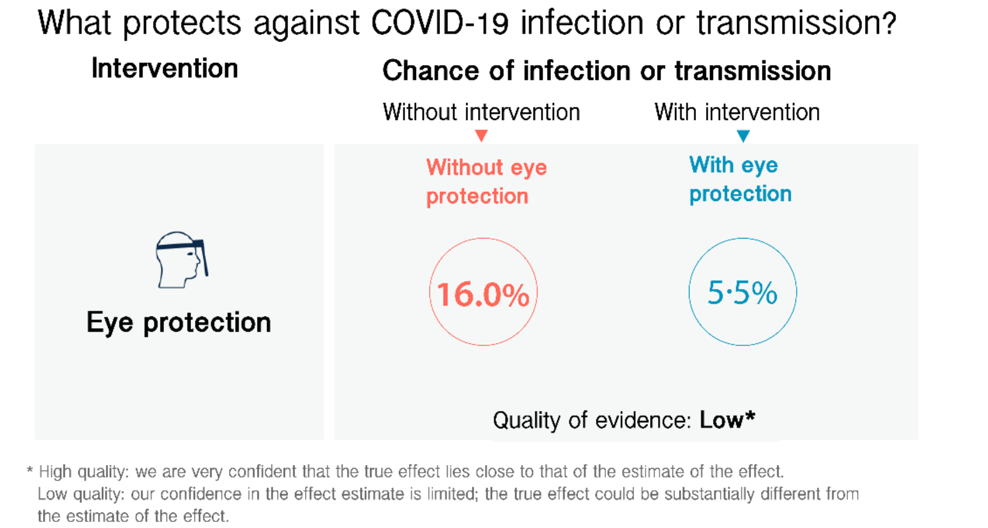

Low quality of evidence – percentages and icon arrays (n = 198):

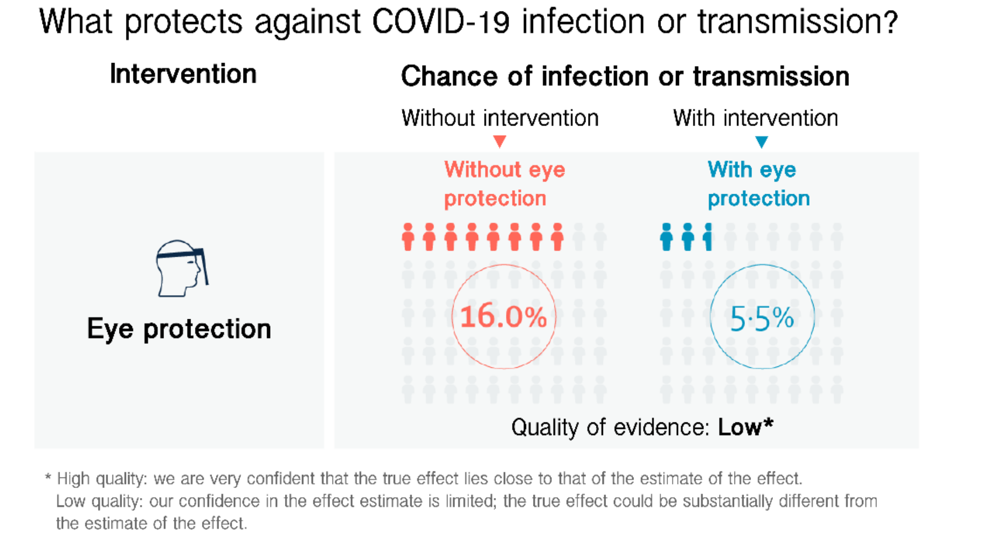

No quality of evidence – percentages only (n = 191):

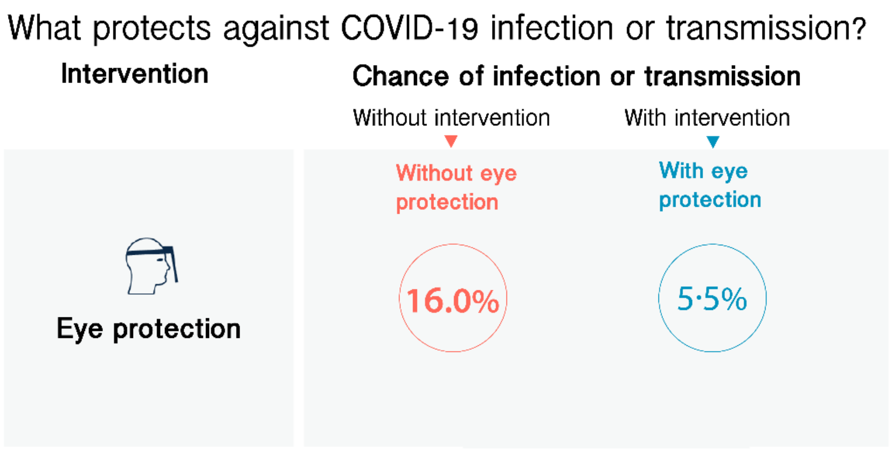

No quality of evidence – percentages and icon arrays (n = 200):

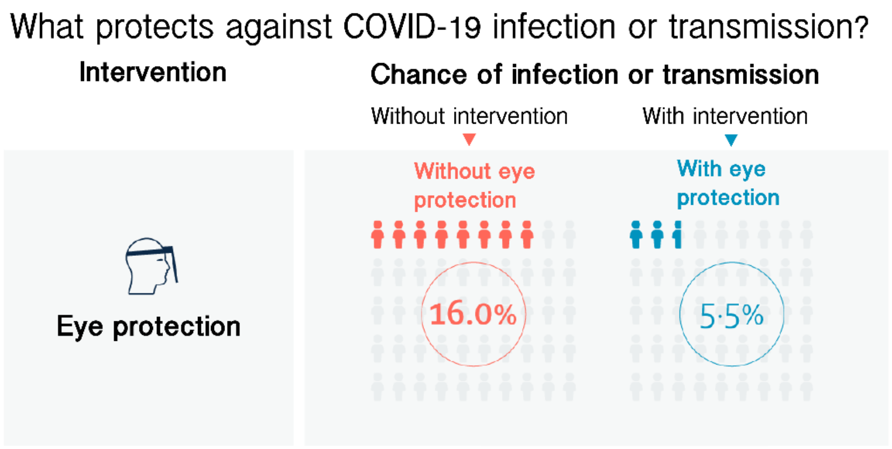

#### Survey measures

**Dependent measures:**
Perceived trustworthiness index items ($\alpha = 0.96$):
How accurate do you think the information on the effectiveness of eye protection is? Not accurate at all (1) - Very accurate (7)
How reliable do you think the information on the effectiveness of eye protection is? Not reliable at all (1) - Very reliable (7)
How trustworthy do you think the information on the effectiveness of eye protection is? Not trustworthy at all (1) - Very trustworthy (7)

Perceived effectiveness:
How effective do you think eye protection is for reducing the chance of infection or transmission of COVID-19? Not effective at all (1) - Very effective (7)

Behavioural uptake:
How likely are you to wear eye protection when in busy public places? Not at all likely (1) - Very likely (7)

**Understanding and effort items:**
Index of ease and completeness of understanding ($r = 0.74$, $rho = 0.75$):
How easy or difficult did you find the information on the effectiveness of eye protection to understand? Very difficult (1) - Very easy (7)
How completely do you feel you understood the information on the effectiveness of eye protection? Not at all (1) - Completely (7)

Effort invested:
How much effort do you feel you had to put into understanding the information on the effectiveness of eye protection? None (1) - A lot (7)

Objective comprehension:
According to the infographic, the chance of infection without the use of eye protection is...
o about twice as high as with eye protection (1)
o less than twice as high as with eye protection (2)
o more than twice as high as with eye protection (3)
o The infographic does not provide this information (4)

#### Model diagnostics

Quality of evidence level and format on perceived trustworthiness:

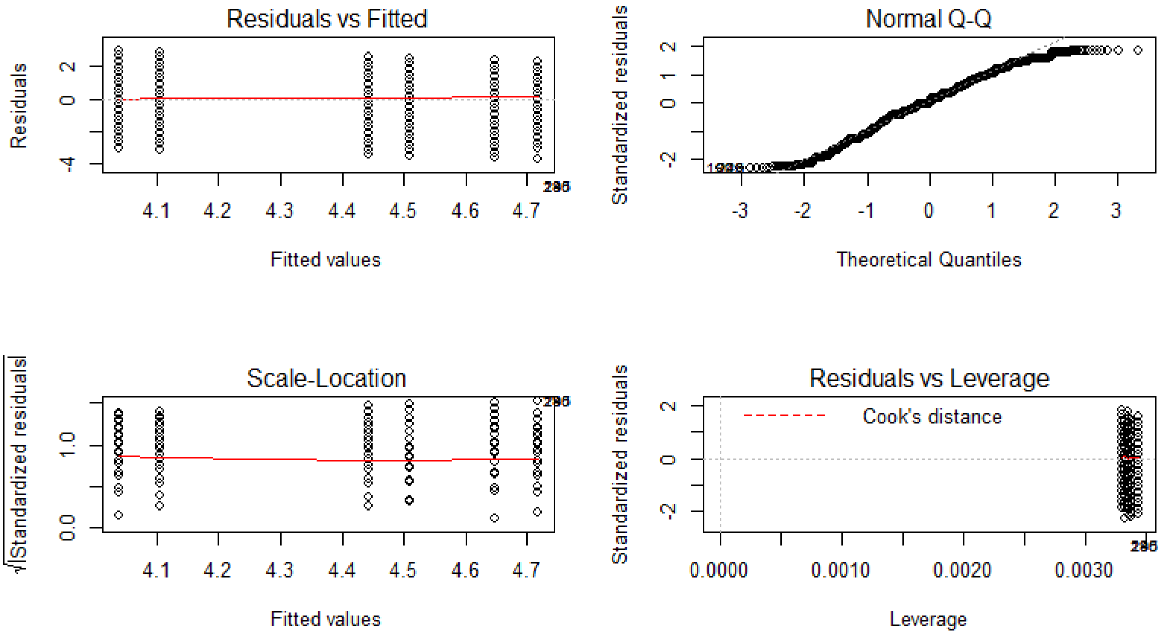

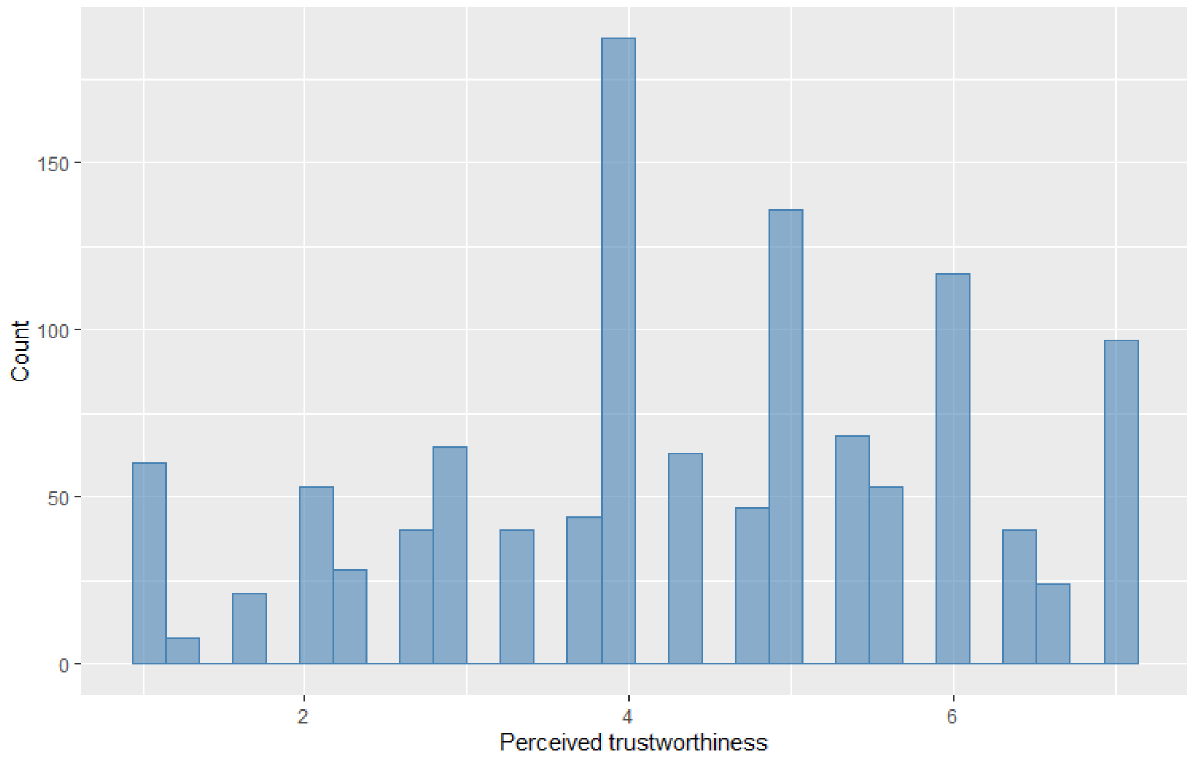

Quality of evidence level and format on perceived effectiveness:

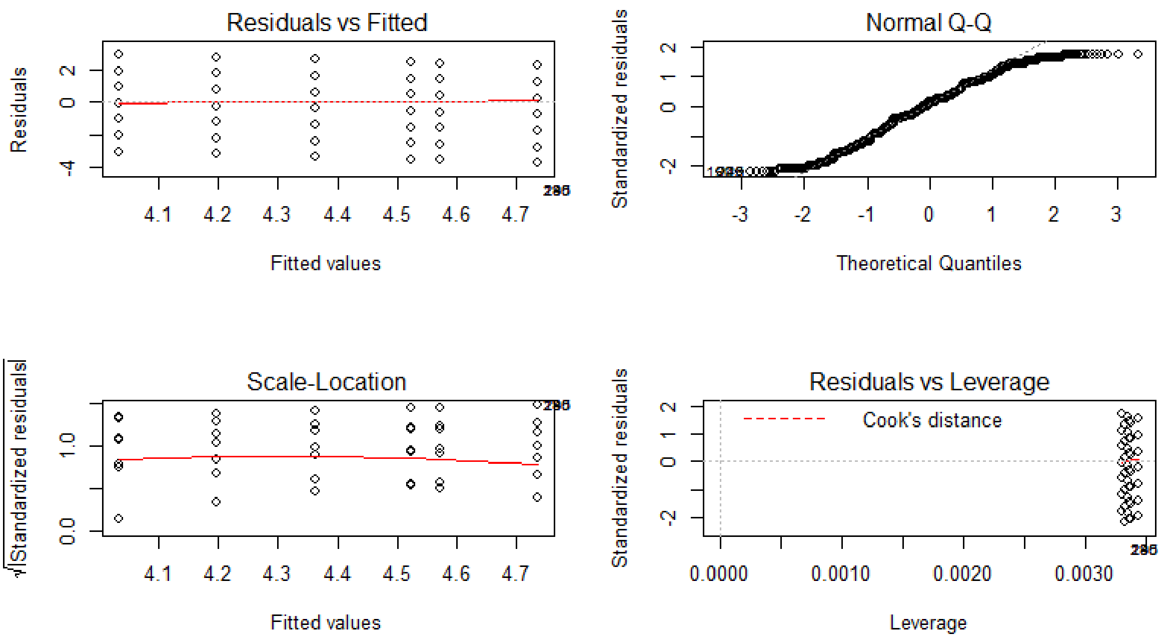

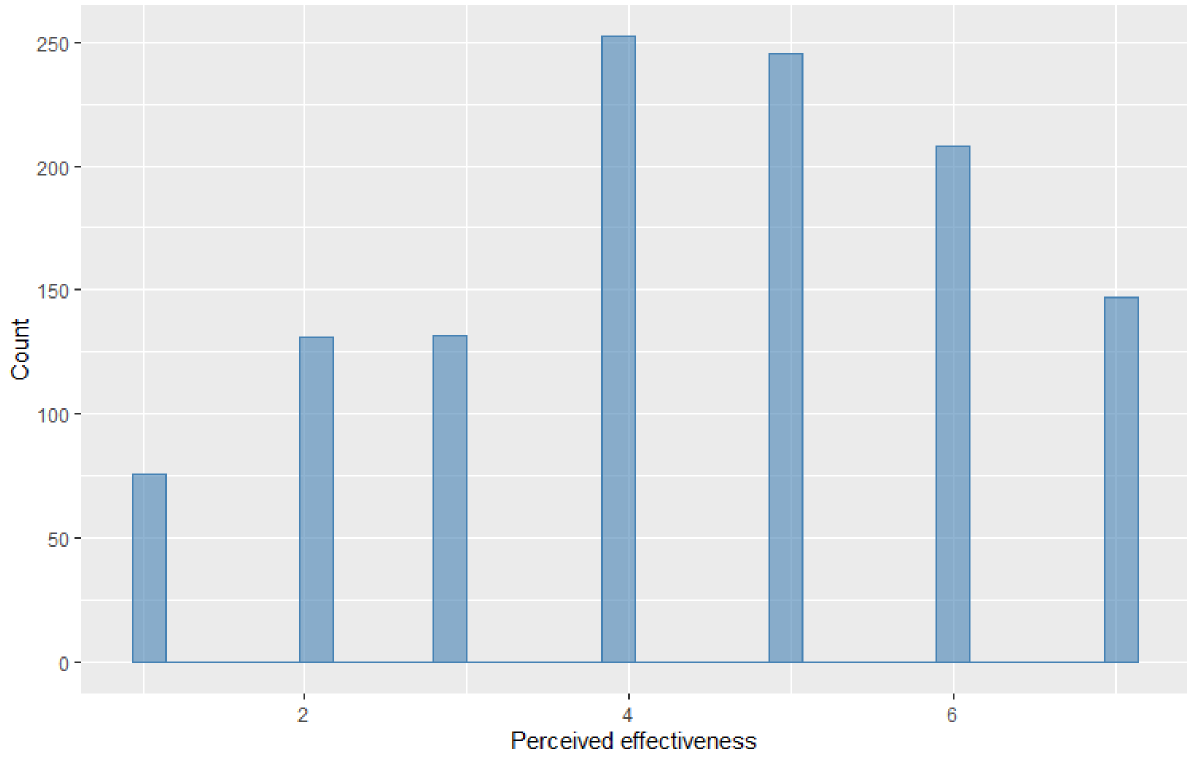

Quality of evidence level and format on behavioural uptake intentions:

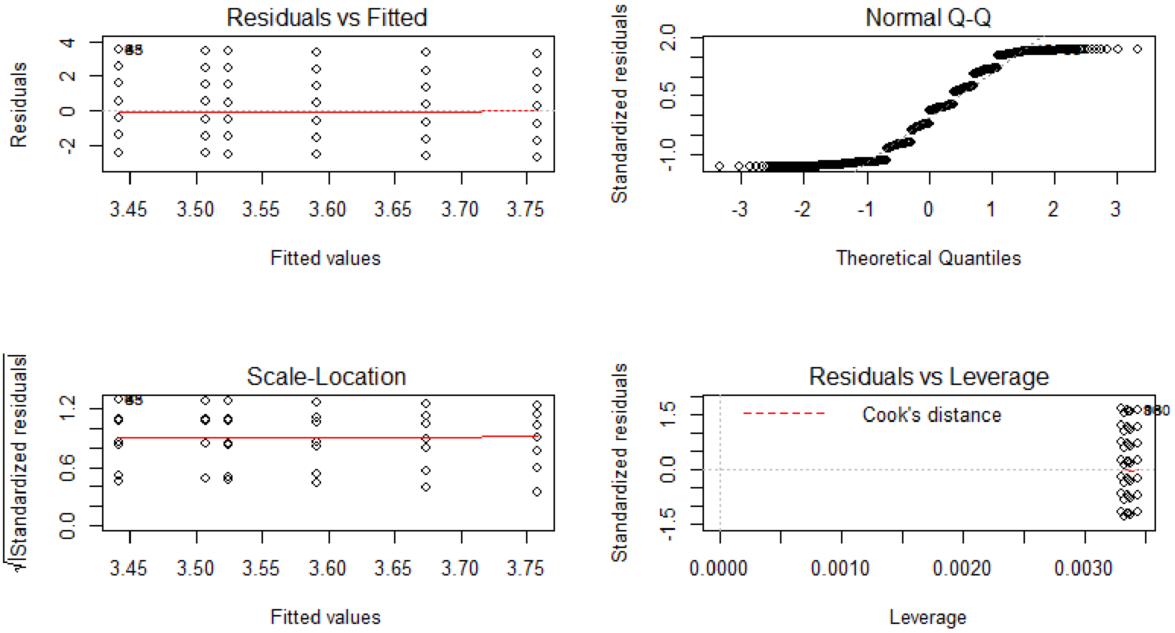

Note: Non-parametric aligned rank transformed ANOVA was conducted, due to skew in the outcome measure.

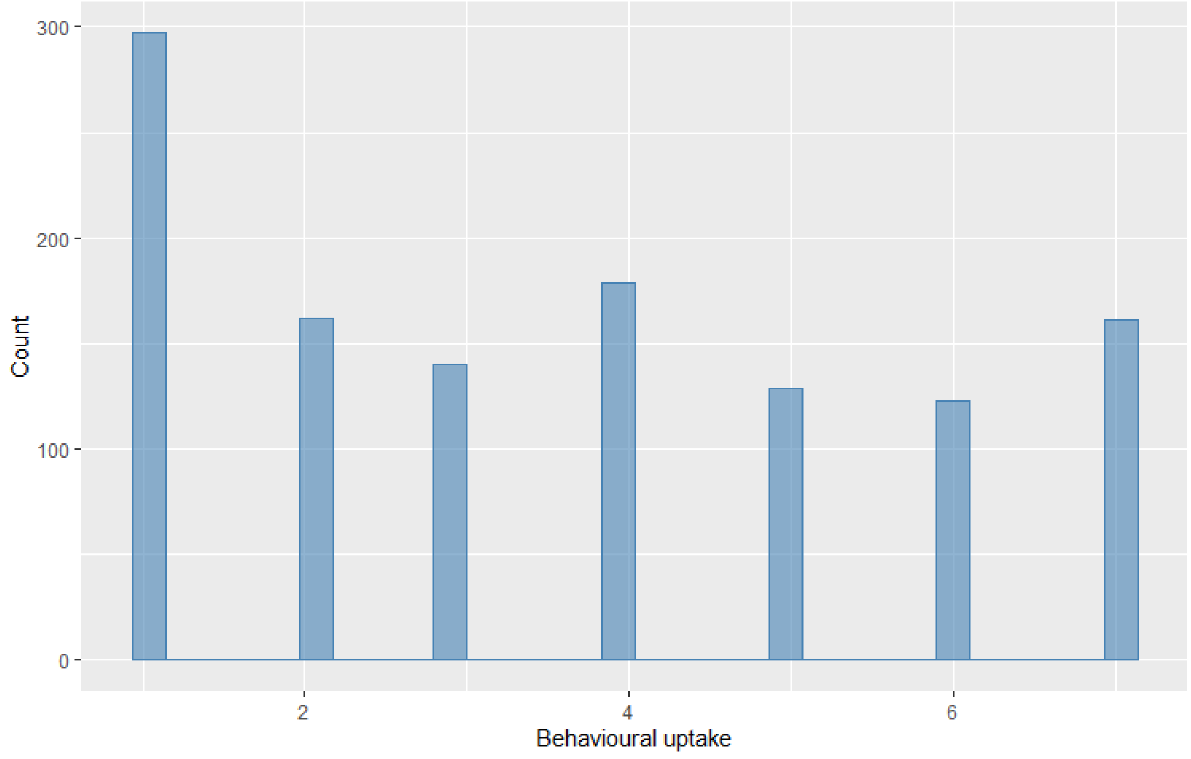

### Supplementary analyses

#### Experiment 1

##### Interaction analysis between quality of evidence level and terminology

We pre-registered to run exploratory interaction analyses between quality of evidence level and wording on our dependent measures. We do not find any significant interactions, neither for perceived trustworthiness ($F(1,945) = 0.03$, $p = .857$), perceived effectiveness ($F(1,945) = 0.21$, $p = .648$), nor for behavioural uptake intentions ($F(1,945) = 0.30$, $p = .585$).

##### The role of priors – effectiveness and quality of evidence views

We had pre-registered to run exploratory interaction analyses between people's priors of the effectiveness of eye protection and the experimentally assigned quality of evidence level, as well as between people's priors of the quality of evidence level underlying the effectiveness of eye protection and the experimentally assigned quality of evidence level.

We collected two measures in Experiment 1 which aimed at capturing people's priors of the effectiveness of wearing eye protection and the underlying quality of evidence level respectively.

The two measures asked:
*Independent of what we might have told you in this study, how effective do you think eye protection is? (slider scale, Not effective at all – Very effective), and*
*Independent of what we might have told you in this study, how high or low do you think the [quality/certainty] of the evidence underlying the effectiveness of eye protection is? (slider scale, Low – High).*

Please note that these measures were collected at the end of the survey, to not prime people ahead of our experimental manipulations. On the flip side though it is therefore possible that people's answers were influenced by the experimental groups they had been assigned to. For instance we observe that people's perceived effectiveness as an outcome measure of our experimental manipulation is highly and significantly correlated with their views on the effectiveness of eye protection supposedly prior of being shown our experimental information ($r = 0.81$, $t(947) = 42.61$, $p < .001$; Spearman's rank correlation rho = 0.80, $p < .001$). We thus advise to interpret the results of our 'priors' analysis with caution, and encourage future research to measure priors ahead of time for a more reliable picture. We report the analyses here nevertheless, in order to satisfy the pre-registration.

We do not find any significant interactions between people's effectiveness priors and the assigned experimental quality of evidence level on perceived trustworthiness ($F(1,945) = 3.60$, $p = .058$), perceived effectiveness ($F(1,945) = 0.09$, $p = .764$), or behavioural uptake intent ($F(1,945) = 0.01$, $p = .914$).

We likewise do not find any significant interactions between people's quality of evidence priors and the assigned experimental quality of evidence level on perceived trustworthiness ($F(1,945) = 1.01$, $p = .315$), perceived effectiveness ($F(1,945) = 0.59$, $p = .442$), nor behavioural uptake intentions ($F(1,945) = 0.02$, $p = .897$).

Our exploratory pre-registered analysis plan furthermore included to control for people's priors of effectiveness and quality of evidence in our models. We run our main effects models presented in the main manuscript on perceived trustworthiness, perceived effectiveness, and behavioural uptake, entering the two priors as covariates. We run separate models for the two priors as they are highly and significantly correlated ($r = 0.86$, $t(947) = 50.82$, $p < .001$; we run non-parametric correlation analysis

as a robustness check since the measures are non-normally distributed: Spearman's rank correlation rho = 0.86, $p < .001$).
The main effect of quality of evidence level for perceived trustworthiness reported in the main manuscript stays significant after controlling for people's effectiveness prior ($F(1,945) = 28.35$, $p < .001$, $\eta_G^2 = 0.029$) as well as people's quality of evidence prior ($F(1,945) = 16.46$, $p < .001$, $\eta_G^2 = 0.017$).
The main effect of quality of evidence level for perceived effectiveness likewise stays significant after controlling for people's effectiveness prior ($F(1,945) = 9.01$, $p = .003$, $\eta_G^2 = 0.009$). When controlling for people's quality of evidence prior, the main effect of quality of evidence level does no longer reach significance ($F(1,945) = 1.86$, $p = .173$). Likewise, the main effect of quality of evidence level on behavioural intentions does not longer reach significance controlling for people's effectiveness priors ($F(1,945) = 1.10$, $p = .295$) and quality of evidence priors ($F(1,945) = 0.01$, $p = .933$).

#### Self-reported (in-)congruency between priors and presented info

For exploratory purposes we included two measures of self-reported match between people's priors of the effectiveness of eye protection and the effectiveness level shown in the infographic, as well as between people's priors of the underlying quality of evidence level and the quality of evidence level indicated in the infographic.

The two measures were:

*To what extent did the effectiveness of eye protection given in the infographic match what you thought it was? I thought eye protection was much less effective (1) - I thought eye protection was about as effective as shown in the infographic (4) - I thought eye protection was much more effective (7)*

*To what extent did the [quality/certainty] of evidence level underlying the effectiveness of eye protection as shown in the infographic match what you thought it was? I thought the [quality/certainty] of evidence level was much lower (1) - I thought the [quality/certainty] of evidence level was about the same as in the infographic (4) - I thought the [quality/certainty] of evidence level was much higher (7)*

The distributions of both items indicated that most people had thought that eye protection was as effective as shown to them in the infographic, and that the underlying quality of evidence level was the same as shown to them in the infographic.

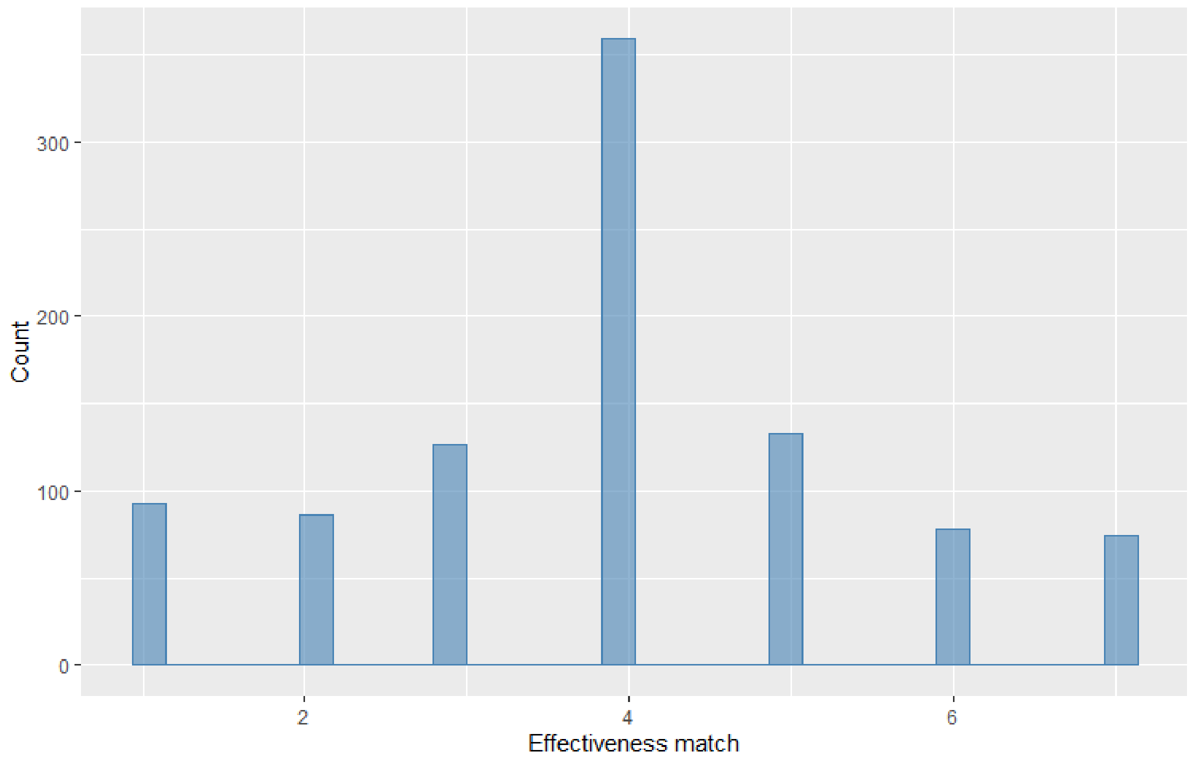

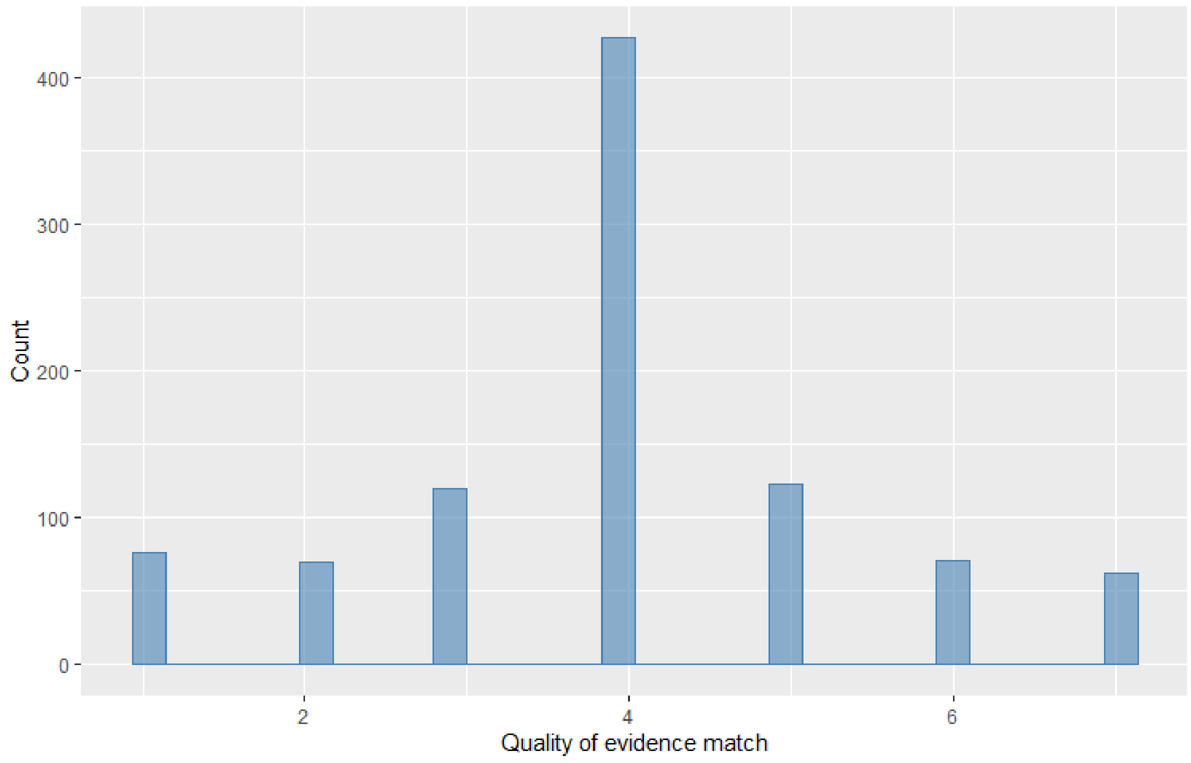

While for effectiveness this could very well be the case, and would point towards well-informed participants, the findings for quality of evidence suggests that people's responses were influenced by the experimental manipulation (these measures were collected at the end of the survey). The distributions of the quality of evidence priors match for both the high and the low quality of evidence experimental groups, both show the same pattern with a large percentage of the participants indicating a match. These trends highlight that people might not be able to or might not want to report on their priors after having received information. It is thus difficult to tell from the measures as we had implemented them in this study whether their prior perceptions of the effectiveness levels of eye protection and underlying quality of evidence were in fact congruent with the presented information or not. We encourage further research that measures priors in advance of any experimental manipulation.

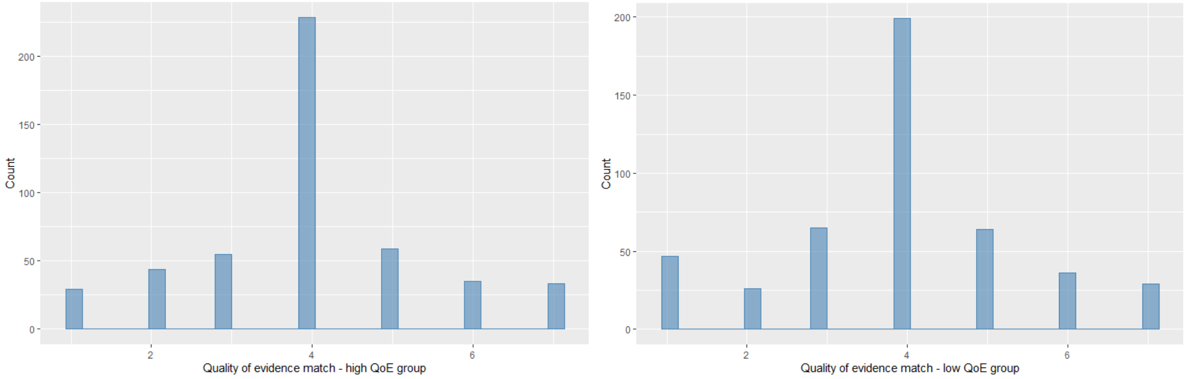

#### Self-reported shift in trust and behavioural intentions

For exploratory purposes we included two measures of people's self-reported shift in trust and behavioural intentions due to the infographic.

The two measures were:

*To what extent has the information in the infographic influenced your trust in the effectiveness of eye protection? It has made me trust it much less (1) - It hasn't changed my trust (4) - It has made me trust it much more (7)*

*How much more or less likely are you to wear eye protection as a result of the information in the infographic? A lot less likely (1) - The information has not changed anything (4) - A lot more likely (7)*

The distributions of both items show that most people indicated that the infographic did not have an influence on their trust and behaviour.

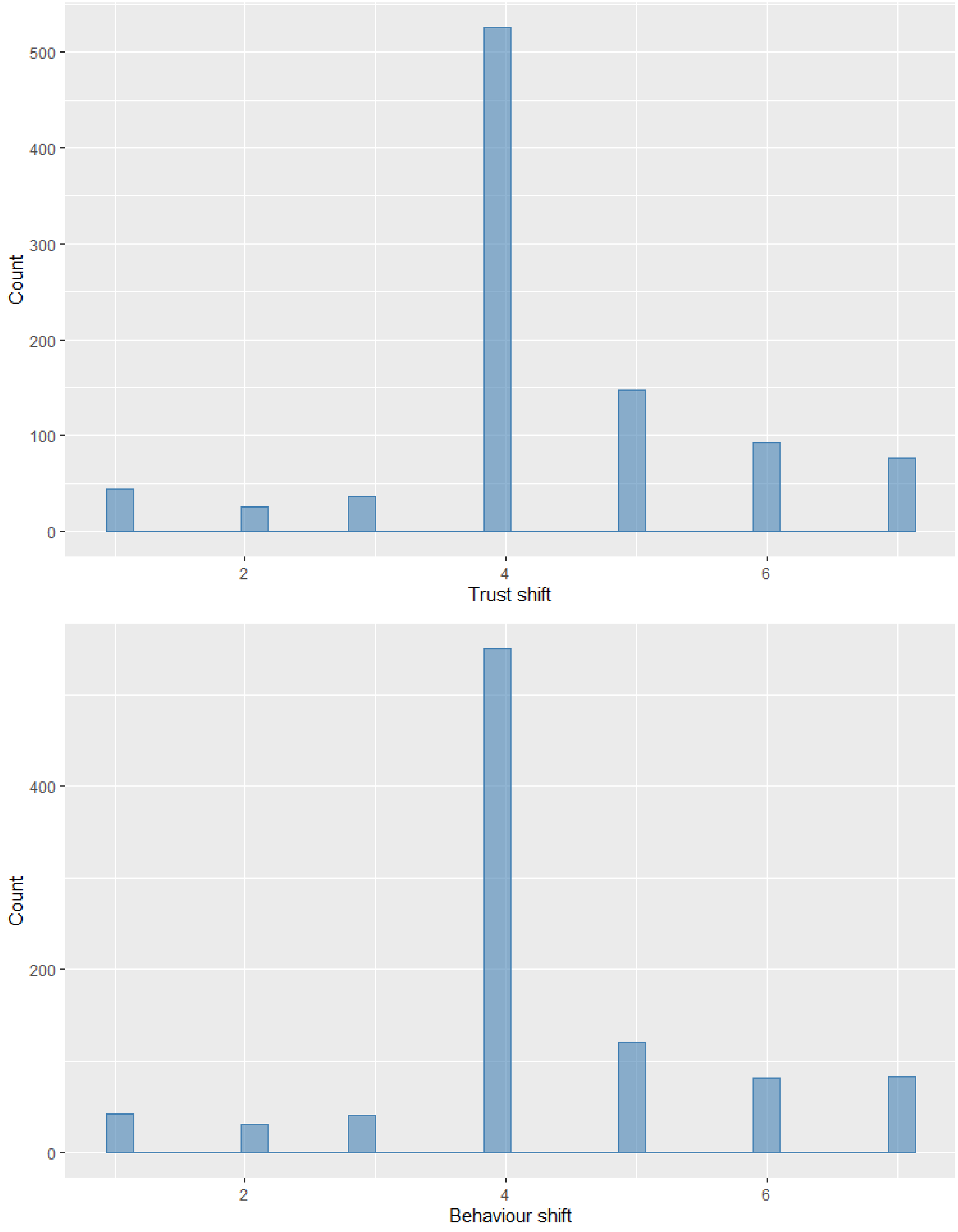

The same descriptive pattern emerges when splitting by those who were in the high quality of evidence group and those who were in the low quality of evidence group.

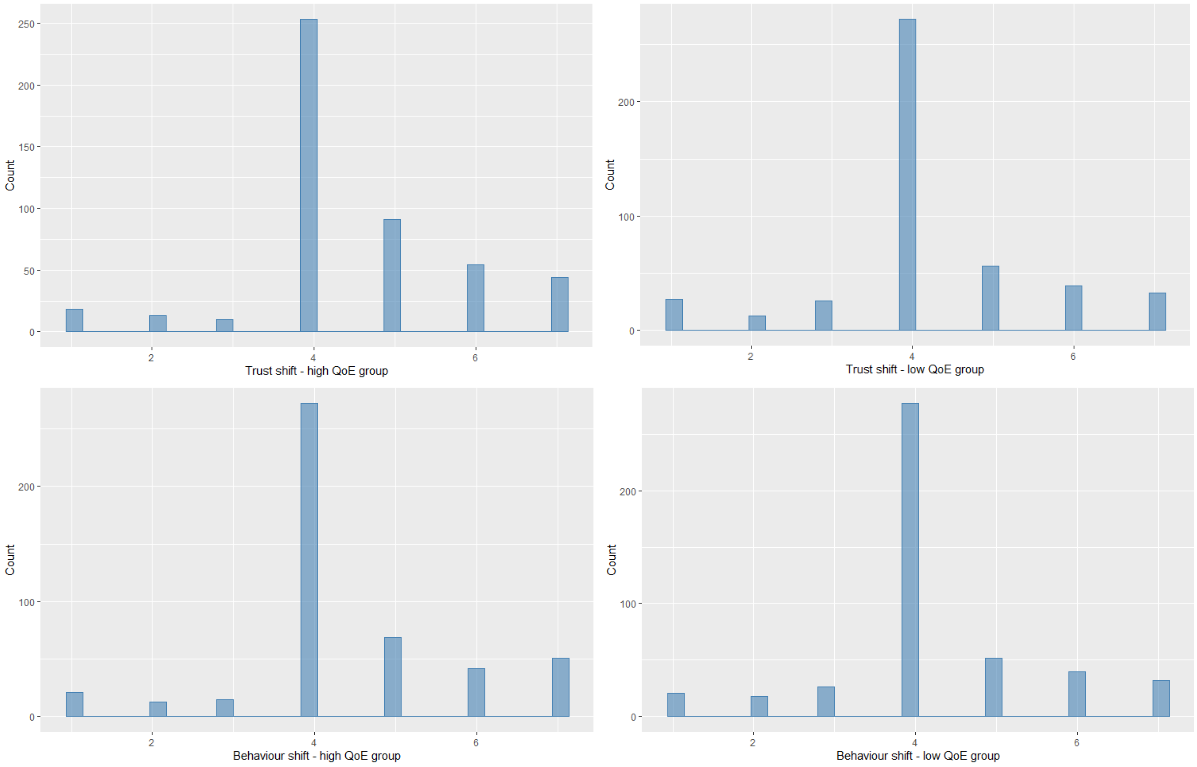

This pattern of the bulk of the responses being "It hasn't changed my trust"/" The information has not changed anything" suggests that to an extent people might not be aware of the influence that the infographic has on them, or that they might not want to admit any influence.
Nevertheless, comparing group averages on reported trust and behavioural shifts due to the infographic, reveals a significant different between the high and low quality of evidence groups. In line with our main experimental findings presented in the main manuscript, people in the high quality of evidence group (*M* = 4.50, 95% CI [4.38,4.61]) report that the infographic made them trust it more compared to those in the low quality of evidence group (*M* = 4.21,95% CI [4.09,4.33], $d_{adj} = 0.22$, OR = 1.49, *t*(944.15) = 3.35, p < .001). We run a non-parametric Mann-Whitney test for robustness purposes. Results are in line with the parametric findings (W= 127728, p < .001). We find the same results comparing the high and low quality of evidence groups on reported behavioural shift. People in the high quality of evidence group (*M* = 4.42, 95% CI [4.30, 4.54]) report that the information in the infographic made them more likely to wear eye protection compared to people in the low quality of evidence group (*M* = 4.22,95% CI [4.10,4.34], $d_{adj} = 0.15$, OR = 1.31, *t*(946.91) = 2.32, p = .021, Mann-Whitney: W = 122473, p = .009).

#### Analysis of exploratory dependent measures

**<u>Trust in the producers of the information</u>**

We also asked participants to indicate how trustworthy they thought the people responsible for producing the infographic on the effectiveness of eye protection are using the following item:

*How trustworthy do you think the people who are responsible for producing the infographic on the effectiveness of eye protection are? Not trustworthy at all (1) - Very trustworthy (7)*

Results showed a main effect of quality of evidence level (F(1,946) = 8.42, p = .004, $\eta_G^2 = 0.009$), such that participants in the low quality of evidence group (*M* = 4.31, 95% CI [4.17,4.46]) indicated significantly lower levels of trust in the producers of the information compared to participants in the high quality of evidence group (*M* = 4.61, 95% CI [4.47,4.74], p = .004, $d_{adj}$ = 0.19, OR = 1.41). No effect of quality wording was observed (F(1,946) = 0.001, p = .976).

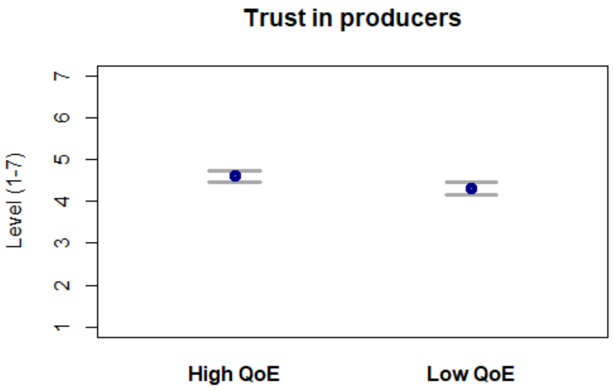

Note: Figure depicts means and 95% confidence intervals for trust in producers for the high quality of evidence group (High QoE) and the low quality of evidence group (Low QoE).

**Support of government policies to enforce wearing of eye protection**

We also collected a measure that probed participant's' support of a government policy to enforce wearing of eye protection with the following item:

"*To what extent do you think the government should require people to wear eye protection in busy public places?*" (not at all (1) – very much (7)).

No main effects were observed, neither for quality of evidence level (F(1,946) = 2.52, p = .113) nor for wording (F(1,946) = 0.05, p = .824). Aligned rank transformed ANOVAs (conducted as a robustness check due to skew in the outcome variable) confirmed the observed parametric results (quality of evidence level: F(1,945) = 2.86, p = .091; wording: F(1,945) = 0.03, p = .862) .
The distribution for the policy support measure showed that a large amount of participants answered on the extreme bottom end of the scale, i.e. not at all supporting eye protection enforcement. This skew in the data rendered this measure somewhat uninformative.

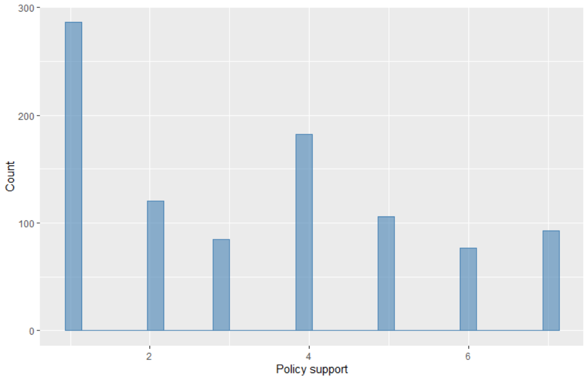

#### Potential moderators – numeracy, prosocial attitudes, and political orientation

We ran several exploratory analyses investigating the role of potential moderators.

We collected a range of additional variables for exploratory analyses. Given that our study dealt with numeric information (percentages on the chance of infection), we investigated the potential effects of numeracy, i.e. whether participants of different numeracy levels would interact differently with the information in the infographic and 'rely' on the quality of evidence information to a different extent.

Previous work had furthermore reported the importance of prosocial attitudes in the context of risk perception and behaviour relating to COVID-19. We thus wanted to test whether prosociality played a role in our study with regards to how people react to the experimental treatments of varying quality of evidence information. Finally, as COVID-19 was a highly politicized topic in the United States at the time of data collection[1], we furthermore tested for potential moderation effects of political orientation.

**Numeracy:**

Numeracy was measured using the sum of the scores of a combination of items. First, participants responded to the adaptive Berlin numeracy test (Cokely et al., 2012). Next, participants completed three items from Schwartz et al. (1997), and finally one single item adapted from Lipkus, Samsa, and Rimer (2001).

The items were:
*Out of 1,000 people in a small town 500 are members of a choir. Out of these 500 members in the choir 100 are men. Out of the 500 inhabitants that are not in the choir 300 are men. What is the probability that a randomly drawn man is a member of the choir?*

*Imagine we are throwing a five-sided die 50 times. On average, out of these 50 throws how many times would this five-sided die show an odd number (1, 3 or 5)?*

*Imagine we are throwing a loaded die (6 sides) 70 times. The probability that the die shows a 6 is twice as high as the probability of each of the other numbers. On average, out of these 70 throws how many times would the die show the number 6?*

*In a forest 20% of mushrooms are red, 50% brown and 30% white. A red mushroom is poisonous with a probability of 20%. A mushroom that is not red is poisonous with a probability of 5%. What is the probability that a poisonous mushroom in the forest is red?*

*Which represents the highest risk of something happening?*

*1 in 100; 1 in 1000; 1 in 10*

*Imagine that we flip a fair coin 1,000 times. What is your best guess about how many times the coin would come up heads in 1,000 flips?*

*In a scratch card lottery, the chance of winning a £10 prize on the card is 1%. What is your best guess about how many people would win a £10 prize if 1,000 people each buy a single scratch card?*

*At a raffle, the chance of winning a car is 1 in 1,000. What percentage of tickets in the raffle win a car?*

Cokely ET, Galesic M, Schulz E, Ghazal S, Garcia-Retamero R. 2012 Measuring Risk Literacy: The Berlin Numeracy Test. *Judgm. Decis. Mak.* **7**, 25–47.
Schwartz LML, Woloshin SS, Black WCW, Welch HGH. 1997 The role of numeracy in understanding the

[1] https://www.journalism.org/2020/10/07/before-trump-tested-positive-for-coronavirus-republicans-attention-to-pandemic-had-sharply-declined/
Graham, A., Cullen, F. T., Pickett, J. T., Jonson, C. L., Haner, M., & Sloan, M. M. (2020). Faith in Trump, moral foundations, and social distancing Defiance during the coronavirus pandemic. *Socius*, *6*, 2378023120956815. https://journals.sagepub.com/doi/full/10.1177/2378023120956815
Grossman, G., Kim, S., Rexer, J. M., & Thirumurthy, H. (2020). Political partisanship influences behavioral responses to governors' recommendations for COVID-19 prevention in the United States. *Proceedings of the National Academy of Sciences*, *117*(39), 24144-24153. https://www.pnas.org/content/117/39/24144.short

benefit of screening mammography. *Ann. Intern. Med.* **127**, 966–972.
Lipkus IM, Samsa G, Rimer BK. 2001 General performance on a numeracy scale among highly educated samples. *Med. Decis. Making* **21**, 37–44. (doi:10.1177/0272989X0102100105)

We find a small significant interaction between numeracy and quality of evidence level for perceived trustworthiness (F(1,943) = 10.18, p = .001, $\eta_G^2$ = 0.011), i.e. the effect of quality of evidence level on perceived trustworthiness depends to an extent on numeracy. For lower numeracy levels the effect of quality of evidence is less pronounced compared to higher numeracy levels. The higher the numeracy the more pronounced the experimental effect of quality of evidence, i.e. lower trust for low quality of evidence information and higher trust for high quality of evidence information. The difference in perceived trustworthiness between low and high quality of evidence gets smaller the lower the numeracy. This could hint to different levels of engagement with or understanding of the information in the infographic for people with lower numeracy compared to higher numeracy.
We find the same pattern for perceived effectiveness. There is a significant interaction between quality of evidence level and numeracy (F(1,943) = 3.89, p = .049, $\eta_G^2$ = 0.004) in the same direction as for trustworthiness.
We do not find a significant interaction for intentions to wear eye protection (F(1,943) = 1.50, p = .221).

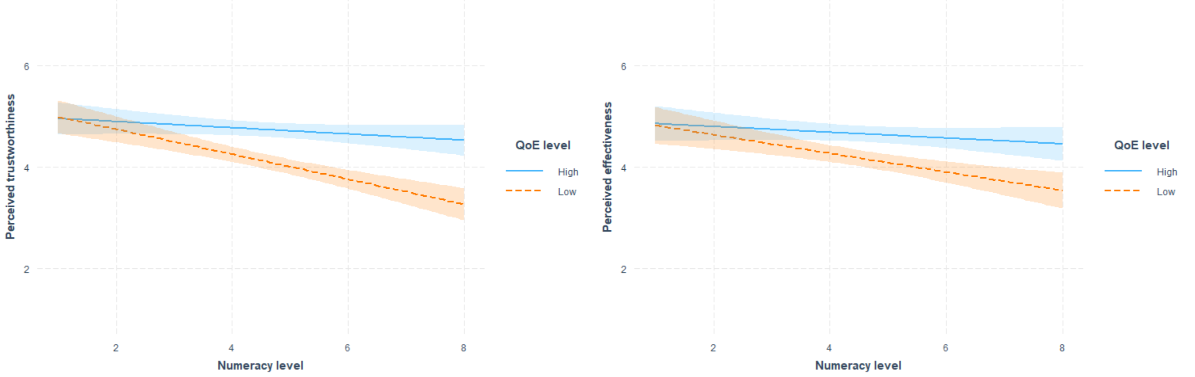

Note: Interaction plots of quality of evidence level and numeracy on perceived trustworthiness (left) and perceived effectiveness (right). Light shading around trend lines denotes 95% confidence band.

**Prosociality:**

Prosociality was measured with a single item:
*To what extent do you think it's important to do things for the benefit of others and society, even if they have some costs to you personally? (7 point Likert scale, 'not at all' – 'very much so')*

We find a small significant interaction between people's prosocial tendencies and quality of evidence level for perceived trustworthiness (F(1,945) = 5.51, p = .019, $\eta_G^2$ = 0.006), such that for higher prosociality levels the effect of receiving high versus low quality of evidence information is more pronounced compared to lower prosociality levels.
One potential explanation could that the more prosocial people are the more they engage with the quality of evidence information, such that this higher engagement may lead to the expression of the differential effect of high versus low quality of evidence on trust, i.e. higher trust for high quality of evidence compared to low quality of evidence. For people low on prosocial tendencies the quality of evidence level they are presented with makes less of a difference for their trust, possible because they engage less with the information.
No significant interactions emerged for perceived effectiveness (F(1,945) = 0.68, p = .409) and behavioural intentions (F(1,945) = 1.59, p = .207).

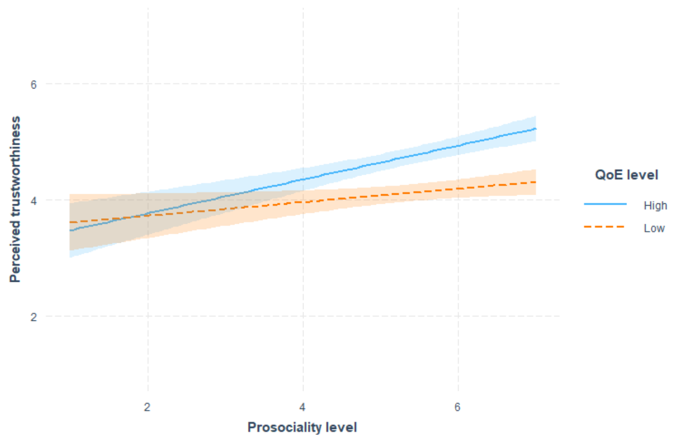

Note: Interaction plot of quality of evidence level and prosociality on perceived trustworthiness. Light shading around trend lines denotes 95% confidence band.

**Political orientation:**

We measured political orientation with the single item:

*Where do you feel your political views lie on a spectrum of left wing (or liberal) to right wing (or conservative)? (7 point Likert scale, 'very left wing/liberal' to 'very right wing/conservative')*

A significant interaction between political orientation and quality of evidence level emerged for perceived trustworthiness ($F(1,945) = 5.78$, $p = .016$, $\eta_G^2 = 0.006$), such that for more right wing/conservative leaning participants the effect of quality of evidence (i.e. higher trust for high quality of evidence and lower trust for low quality of evidence) is less pronounced compared to more left wing/liberal leaning participants.
No significant interactions emerged for perceived effectiveness ($F(1,945) = 2.03$, $p = .154$) or behavioural uptake intent ($F(1,945) = 0.23$, $p = .634$).

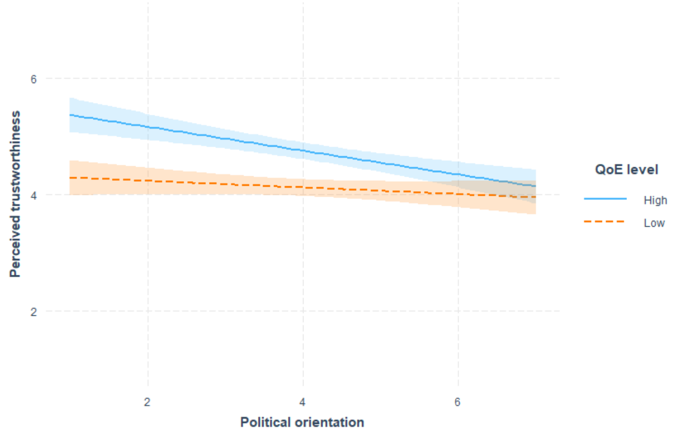

Note: Interaction plot of quality of evidence level and political orientation on perceived trustworthiness. Light shading around trend lines denotes 95% confidence band.

#### Experiment 2

##### The role of understanding of the information in the infographic

As detailed in the pre-registration we wanted to explore the influence of understanding on our dependent measures, and especially its role in shaping the effects that quality of evidence information has on trust, perceived effectiveness and behaviour.
We therefore pre-registered to test for an interaction between quality of evidence level and experimentally manipulated format (with the icon array addition supposedly making the effectiveness information easier to understand).

We do not find any interactions between format and quality of evidence level on none of our dependent variables, perceived trustworthiness ($F(2,1185) = 0.35$, $p = .703$), perceived effectiveness ($F(2,1185) = 0.04$, $p = .957$), and behavioural uptake ($F(2,1185) = 0.39$, $p = .675$)[2].

We furthermore pre-registered to test for an interaction through self-reported measures of understanding; such as ease and completeness of understanding, effort invested in understanding, and objective understanding of the numeric information in the infographic.

The following measures were used:
*Ease and completeness of understanding:*
*How easy or difficult did you find the information on the effectiveness of eye protection to understand? Very difficult (1) - Very easy (7)*
*How completely do you feel you understood the information on the effectiveness of eye protection? Not at all (1) - Completely (7)*

Effort invested:
*How much effort do you feel you had to put into understanding the information on the effectiveness of eye protection? None (1) - A lot (7)*

Objective comprehension:
*According to the infographic, the chance of infection without the use of eye protection is...*
o *about twice as high as with eye protection (1)*
o *less than twice as high as with eye protection (2)*
o *more than twice as high as with eye protection (3)*
o *The infographic does not provide this information (4)*

We do not find an interaction between self-reported ease and completeness of understanding of the infographic and quality of evidence level on trustworthiness of the information ($F(2,1185) = 1.69$, $p = .185$), perceived effectiveness ($F(2,1185) = 2.93$, $p = .054$), or behavioural uptake intentions ($F(2,1185) = 0.20$, $p = .819$).
For our measure of objective understanding of the effectiveness information given in the infographic, we likewise do not find any significant interactions on perceived trustworthiness ($F(2,1185) = 1.68$, $p = .188$), perceived effectiveness ($F(2,1185) = 1.42$, $p = .241$), or behavioural uptake intentions ($F(2,1185) = 0.22$, $p = .806$). Finally, for effort invested we do not find an interaction with quality of evidence level on neither trustworthiness ($F(2,1185) = 0.38$, $p = .688$), perceived effectiveness ($F(2,1185) = 1.57$, $p = .208$), nor behaviour ($F(2,1185) = 0.09$, $p = .914$).[3]

---

[2] Given the skew in the behavioural uptake outcome measure, we also performed non-parametric aligned rank transformed ANOVA as a robustness check. No significant interaction was observed ($F(2,1185) = 0.90$, $p = .407$).
[3] As we had noted in our pre-registration that we were in particular interested in the interaction between effort invested and our dependent outcomes for the high versus low quality of evidence contrast, we ran another set of interaction analyses, using only a subset of our data consisting of only the high and low quality of evidence groups. Results were in line with the analyses presented for the whole dataset; i.e. no significant interactions emerged.

For invested effort we furthermore pre-registered to explore main effects on our dependent measures, independent of quality of evidence level.
We did find a main effect of effort invested on trustworthiness independent of quality of evidence level (F(1,1187) = 34.20, p < .001, $\eta_G^2$ = 0.028), indicating that the more effort people invested in understanding the infographic the higher they perceive the trustworthiness of the information (b = 0.14, SE = 0.02, p < .001). We also find main effects in the same direction for perceived effectiveness (F(1,1187) = 36.52, p < .001, $\eta_G^2$ = 0.030) and behavioural uptake (F(1,1187) = 70.77, p < .001, $\eta_G^2$ = 0.056), such that the more effort people invested the higher was their perceived effectiveness of eye protection (b = 0.15, SE = 0.02, p < .001) and the more likely they were to indicate to want to wear eye protection (b = 0.25, SE = 0.03, p < .001).

#### The role of priors – effectiveness and quality of evidence views

As for Experiment 1, we had pre-registered to run exploratory interaction analyses between people's priors of the effectiveness of eye protection and the experimentally assigned quality of evidence level, as well as between people's priors of the quality of evidence level underlying the effectiveness of eye protection and the experimentally assigned quality of evidence level.

We collected two measures which aimed at capturing people's priors of the effectiveness of wearing eye protection and the underlying quality of evidence level respectively.

The two measures asked:

*Independent of what we might have told you in this study, how effective do you think eye protection is? (slider scale, Not effective at all – Very effective).*

*Independent of what we might have told you in this study, how high or low do you think the quality of the evidence underlying the effectiveness of eye protection is? (slider scale, Low – High),* for those experimental groups that had received quality of evidence information in the infographic, and
*How high or low do you think the quality of the evidence underlying the effectiveness of eye protection is? (slider scale, Low-High),* for the control group that had not received any quality of evidence information in the infographic.

As in Experiment 1, these measures were collected at the end of the survey, to not prime people ahead of our experimental manipulations. It is therefore possible that people's answers were influenced by the experimental groups they had been assigned to.

For instance we observe that people's perceived effectiveness as an outcome measure of our experimental manipulation is highly and significantly correlated with their views on the effectiveness of eye protection supposedly prior of being shown our experimental information (r = 0.79, *t*(1189) = 44.11, p < .001; Spearman's rank correlation rho = 0.79, p < .001). We thus advise to interpret the results of our 'priors' analysis with caution, and encourage future research to measure priors ahead of time for a more reliable picture. We report the analyses here nevertheless, in order to satisfy the pre-registration.

We do not find any significant interactions of people's effectiveness priors and the assigned experimental quality of evidence level on perceived trustworthiness (F(2,1185) = 0.78, p = .460), perceived effectiveness (F(2,1185) = 0.03, p = .967), or behavioural uptake intent (F(2,1185) = 0.54, p = .581).

We likewise do not find any significant interactions of people's quality of evidence priors and the assigned experimental quality of evidence level on perceived trustworthiness (F(2,1185) = 0.47, p =

.626), perceived effectiveness ($F(2,1185) = 0.81$, $p = .445$), nor behavioural uptake intentions ($F(2,1185) = 0.37$, $p = .691$).

Our exploratory pre-registered analysis plan furthermore included to control for people's priors of effectiveness and quality of evidence in our models. We test whether our significant main effects reported in the main manuscript stay significant after controlling for people's priors (entering them as covariates). We run separate models for the two priors as they are highly and significantly correlated ($r = 0.82$, $t(1189) = 50.17$, $p < .001$; we ran non-parametric correlation analysis as a robustness check since the measures are non-normally distributed: Spearman's rank correlation rho = 0.83, $p < .001$).
The main effect of quality of evidence level for perceived trustworthiness reported in the main manuscript stays significant after controlling for people's effectiveness prior ($F(2,1186) = 11.90$, $p < .001$, $\eta_G^2 = 0.020$) as well as people's quality of evidence prior ($F(2,1186) = 6.22$, $p = .002$, $\eta_G^2 = 0.010$).
The main effect of quality of evidence level for perceived effectiveness likewise stays significant after controlling for people's effectiveness prior ($F(2,1186) = 7.04$, $p < .001$, $\eta_G^2 = 0.012$). When controlling for people's quality of evidence prior, the main effect of quality of evidence level on perceived effectiveness does no longer reach significance ($F(2,1186) = 2.30$, $p = .101$).

#### Self-reported (in-)congruency between priors and presented info

For exploratory purposes, as in Experiment 1, we included two measures of self-reported match between people's priors on the effectiveness of eye protection and the effectiveness level shown in the infographic, as well as between people's priors on the underlying quality of evidence level and the quality of evidence level indicated in the infographic.

The two measures were:

*To what extent did the effectiveness of eye protection given in the infographic match what you thought it was? I thought eye protection was much less effective (1) - I thought eye protection was about as effective as shown in the infographic (4) - I thought eye protection was much more effective (7)*

*To what extent did the quality of evidence level underlying the effectiveness of eye protection as shown in the infographic match what you thought it was? I thought the quality of evidence level was much lower (1) - I thought the quality of evidence level was about the same as in the infographic (4) - I thought the quality of evidence level was much higher (7)*

The distributions of both items indicated that most people had thought that eye protection was as effective as shown to them in the infographic, and that the underlying quality of evidence level was the same as shown to them in the infographic.

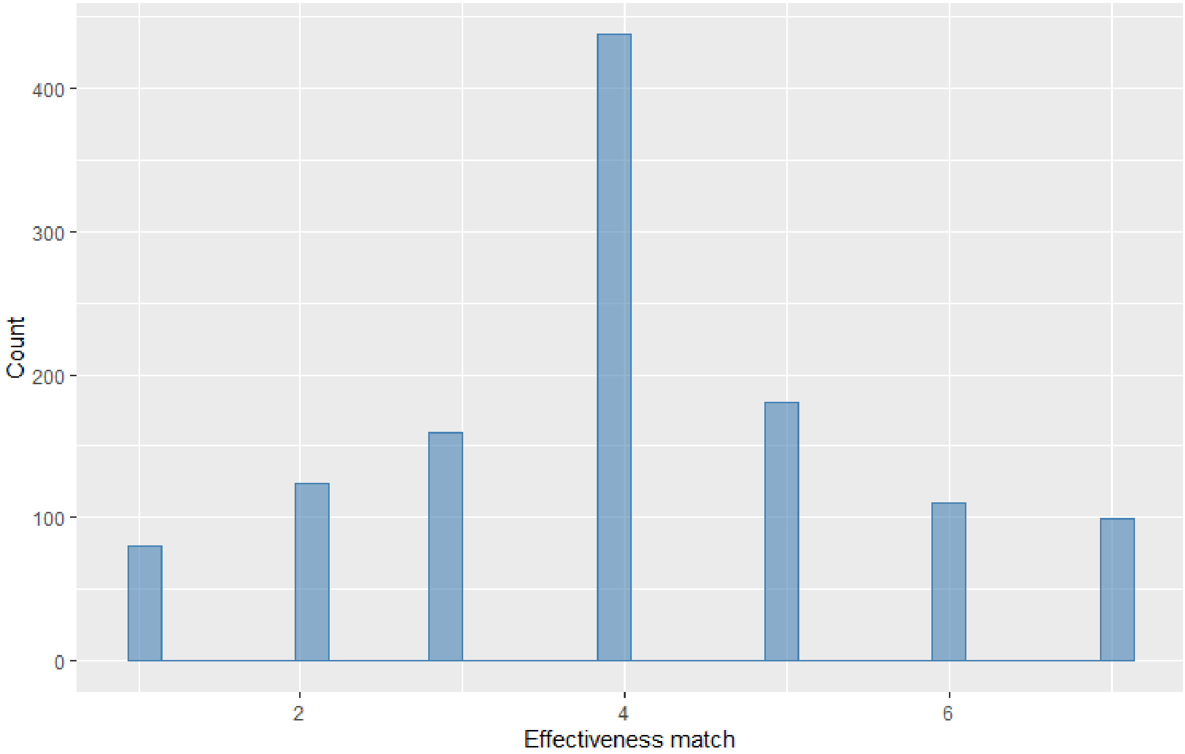

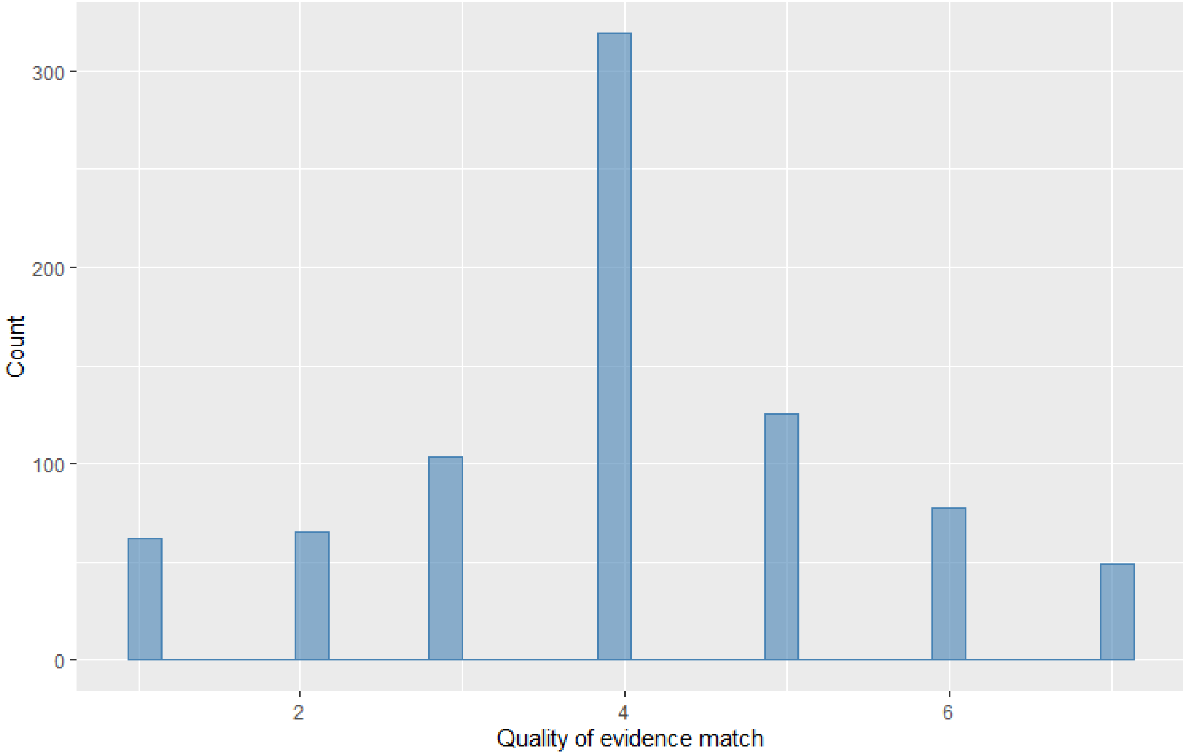

As in Experiment 1 we split the quality of evidence priors by experimental quality of evidence group to investigate whether the high proportion of people indicating an exact match is likely an indication for participants being influenced by the experimental manipulation they had seen.

As in Experiment 1, the distributions for the quality of evidence priors match for both the high and the low quality of evidence experimental groups. Both exhibit the same pattern with participants predominantly indicating a match between their supposed priors and what they were shown. Given random assignment of the treatment groups these results seem to point toward a bias in the measure stemming from the experimental manipulation as outlined above.

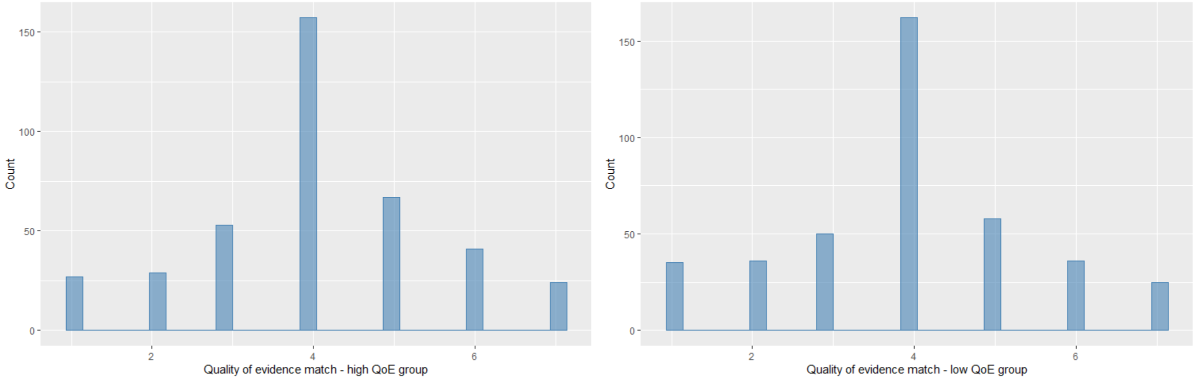

#### Self-reported shift in trust and behavioural intentions

For exploratory purposes we included the same two measures of people's self-reported shift in trust and behavioural intentions due to the infographic as we did in Experiment 1.

The two measures were:

*To what extent has the information in the infographic influenced your trust in the effectiveness of eye protection? It has made me trust it much less (1) - It hasn't changed my trust (4) - It has made me trust it much more (7)*

*How much more or less likely are you to wear eye protection as a result of the information in the infographic? A lot less likely (1) - The information has not changed anything (4) - A lot more likely (7)*

As in Experiment 1, the distributions of both items show that most people indicated that the infographic did not have an influence on their trust and behaviour.

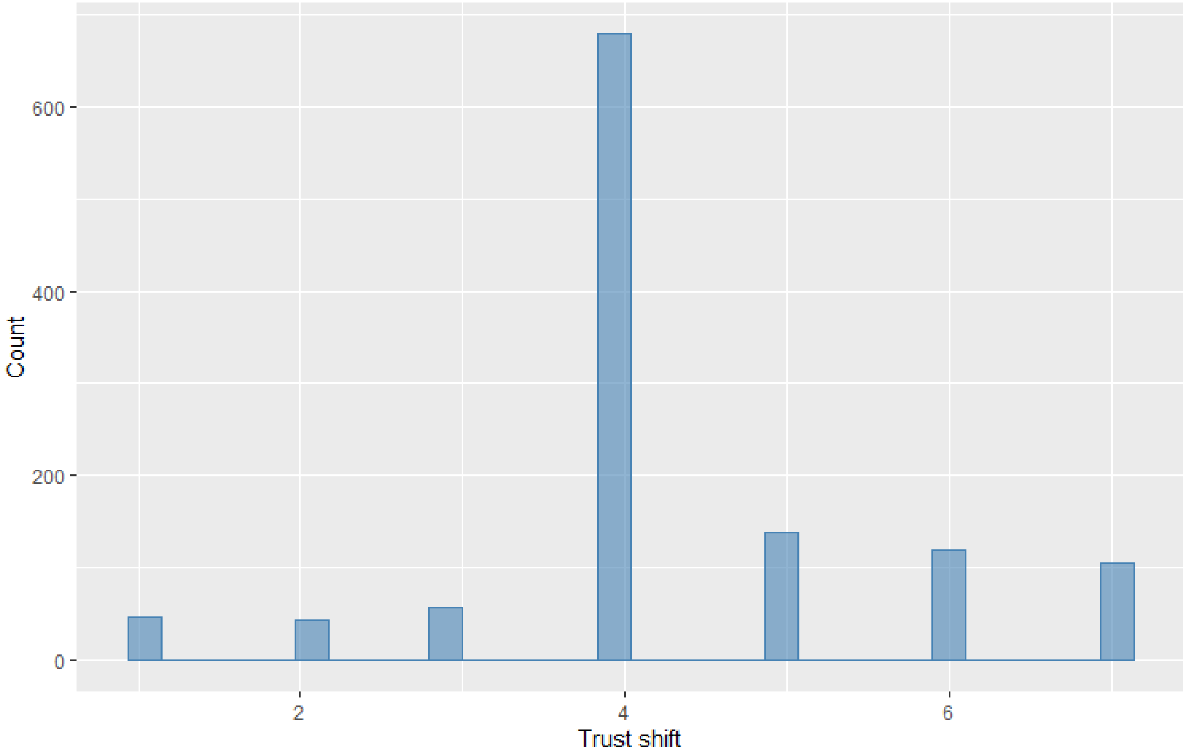

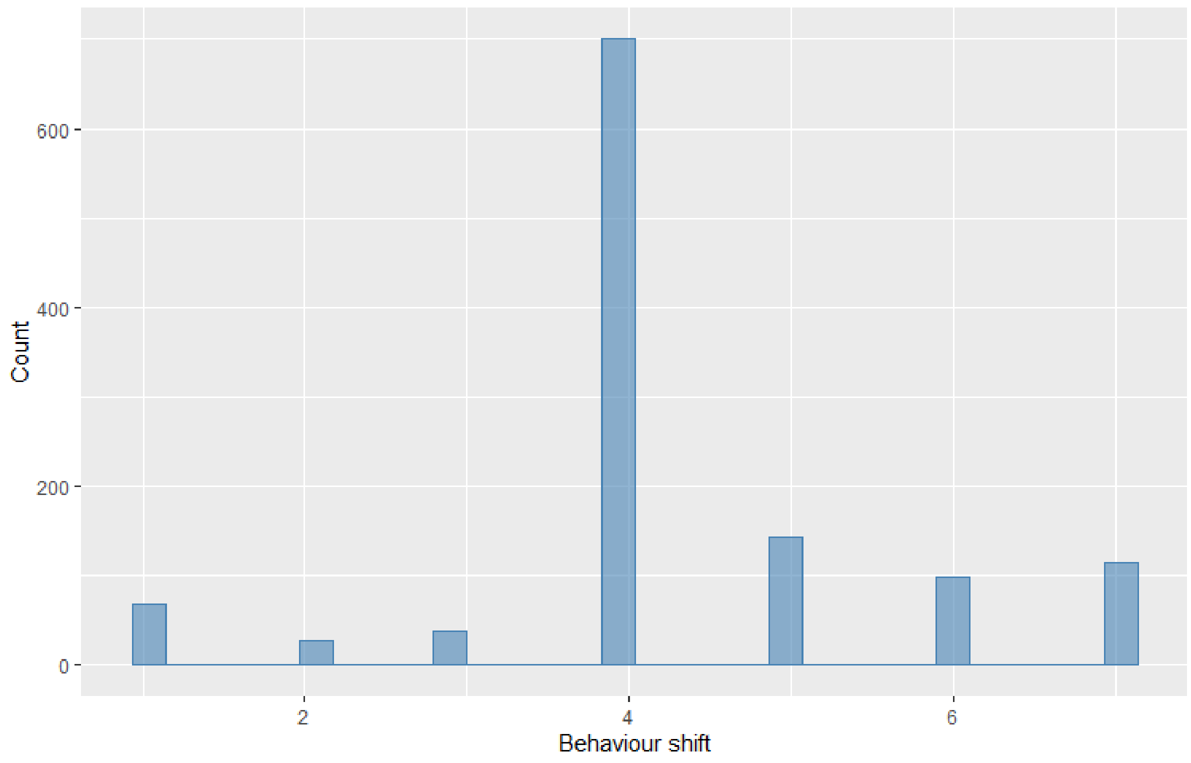

The same descriptive pattern emerges when splitting by those who were in the high quality of evidence, versus low quality of evidence, versus no quality of evidence group.

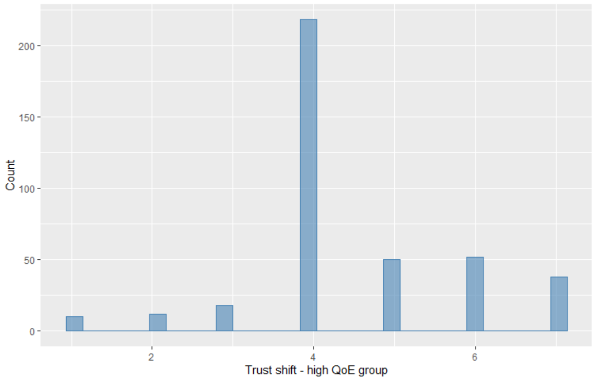

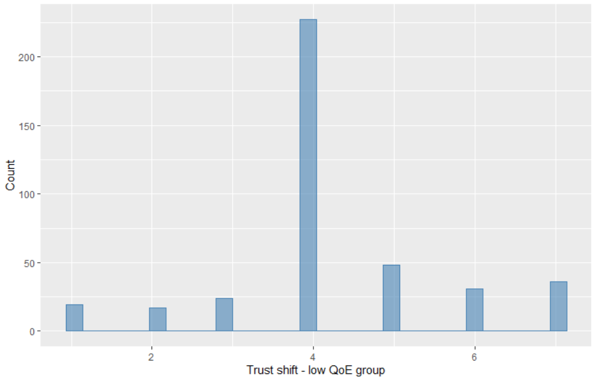

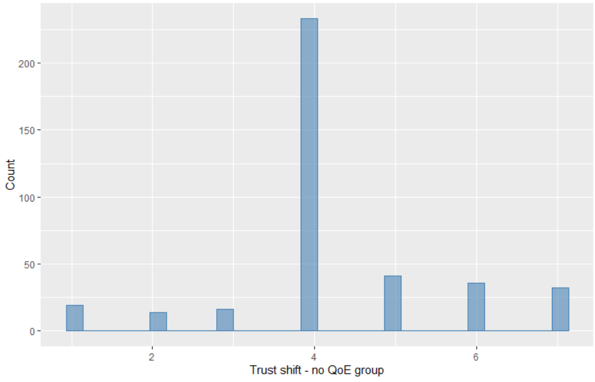

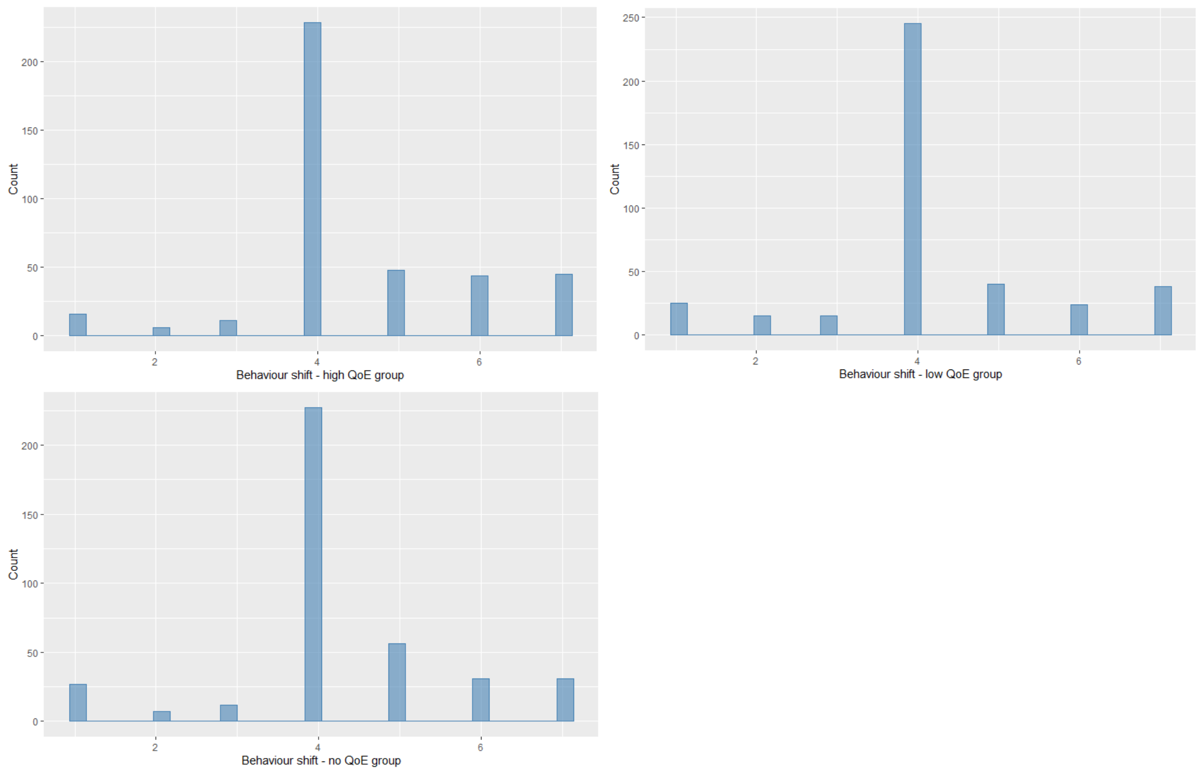

This pattern – which we also observed in Experiment 1 in a similar manner - of the bulk of the responses being "It hasn't changed my trust"/" The information has not changed anything" - suggests that to an extent people might not be aware of the influence that the infographic has on them, or that they might not want to admit any influence.

Nevertheless, we assess differences in reported shift in trust and behaviour by experimental quality of evidence group. A one way Analysis of Variance revealed a significant difference in reported trust shift by quality of evidence levels ($F(2,1188) = 3.90$, $p = .020$, $\eta_G^2 = 0.007$). Post-hoc testing using Tukey HSD correction for multiple comparisons revealed a significant difference between the high quality of evidence group (*M* = 4.49, 95% CI [4.37,4.62]) and the low quality of evidence group (*M* = 4.26, 95% CI [4.12,4.39], $p = .031$, $d_{adj} = 0.18$, OR = 1.39) such that participants in the high quality of evidence group reported a larger shift towards trusting the effectiveness of eye protection more due to the information in the infographic. Of interest is that participants in the low quality of evidence condition still report on average a shift towards more trust rather than less trust (with the mean lying above the neutral scale midpoint). The difference between the high and no quality of evidence group (*M* = 4.28, 95% CI [4.14,4.41]) was not significant ($p = .057$), neither was the difference between the low and no quality of evidence group ($p = .975$). Non-parametric testing for robustness purposes due to the non-normal distribution of the outcome variable likewise revealed a significant group difference in trust shift (Kruskal-Wallis $\chi^2(2) = 7.78$, $p = .020$). Post-hoc comparisons using Mann-Whitney test and Bonferroni adjustment for multiple comparisons ($\alpha$ threshold = 0.05/3 = 0.017) were in line with the parametric results: The difference between the high and low quality of evidence groups was significant ($p = .012$); while the differences between the high and no quality of evidence groups ($p = .024$) and the low and no quality of evidence groups ($p = .775$) were not.
One way Analysis of Variance also revealed a significant difference in reported behaviour shift by quality of evidence levels ($F(2,1188) = 5.39$, $p = .005$, $\eta_G^2 = 0.009$). Post-hoc comparisons showed a significant difference between the high quality of evidence (*M* = 4.50, 95% CI [4.37,4.63]) and the low quality of evidence group (*M* = 4.20, 95% CI [4.07,4.34], $p = .005$, $d_{adj} = 0.22$, OR = 1.49), as well as between the high and the no quality of evidence group (*M* = 4.27, 95% CI [4.13,4.40], $p = .038$, $d_{adj} = 0.18$, OR = 1.39), such that participants in the high quality of evidence group indicated a larger shift in the likelihood of wearing eye protection due to the information in the infographic compared to participants in the low and no quality of evidence groups. There was no significant

difference between the low and no quality of evidence groups (p = .796). Non-parametric analysis showed a significant difference between experimental groups (Kruskal-Wallis $\chi^2(2) = 10.96$, p = .004), in line with the parametric findings. However, post-hoc comparisons using Mann-Whitney tests and Bonferroni adjustment for multiple comparisons (α threshold = 0.05/3 = 0.017) only showed a significant difference between the high and low quality of evidence groups (p = .001), but not for the difference between the high and no quality of evidence groups (p = .040) and the low and no quality of evidence groups (p = .216).

#### Analysis of exploratory dependent measures

##### Trust in the producers of the information

The same item probed people's trust in the producers of the information as used in Experiment 1:

*How trustworthy do you think the people who are responsible for producing the infographic on the effectiveness of eye protection are? Not trustworthy at all (1) - Very trustworthy (7)*

For the question of how trustworthy participants thought the people responsible for producing the infographic on the effectiveness of eye protection are, a main effect of quality of evidence level emerged (F(2,1187) = 3.30, p = .037, $\eta_G^2$ = 0.006). Specifically, the contrast between the high quality of evidence group (*M* = 4.51, 95% CI [4.34,4.67]) and the low quality of evidence group reached significance (*M* = 4.20, 95% CI [4.04,4.37]; p = .027, $d_{adj}$ = 0.18, OR = 1.39), with participants in the high quality of evidence group trusting the producers of the information more compared to participants in the low quality of evidence group. There was no significant difference between the high and no quality of evidence groups (*M* = 4.36, 95% CI [4.19,4.52], p = 0.411), or between the low and no quality of evidence groups (p = 0.399). No effect of format was observed (F(1,1187) = 1.07, p = .300)

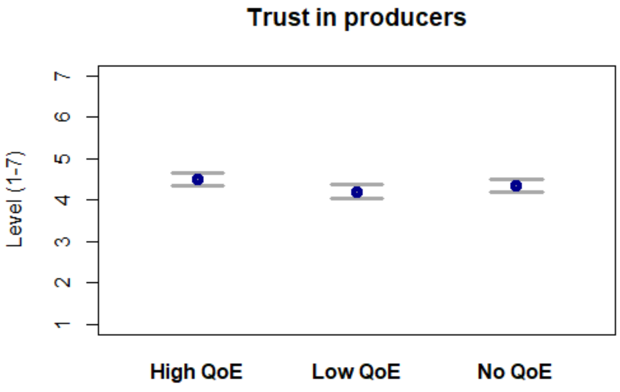

Note: Figure depicts means and 95% confidence intervals for trust in producers for the high quality of evidence group (High QoE), the low quality of evidence group (Low QoE), and the group that did not receive quality of evidence information (No QoE).

##### Support of government policies to enforce wearing of eye protection

As in Experiment 1, the measure that probed participant's' support of a government policy to enforce wearing of eye protection was:

"*To what extent do you think the government should require people to wear eye protection in busy public places?*" (not at all (1) – very much (7)).

No significant differences between experimental groups were observed, neither for quality of evidence level ($F(2,1187) = 2.79$, $p = .062$), nor for format ($F(1,1187) = 1.43$, $p = .233$). Aligned rank transformed ANOVAs (conducted as a robustness check due to skew in the outcome variable) confirmed the observed parametric results (quality of evidence level: $F(2,1185) = 2.94$, $p = .053$, format: $F(1,1185) = 3.18$, $p = .075$).
As in Experiment 1 the distribution for the policy support measure showed that a large amount of participants answered on the extreme bottom end of the scale, i.e. not at all supporting eye protection enforcement. This skew in the data rendered this measure somewhat uninformative.

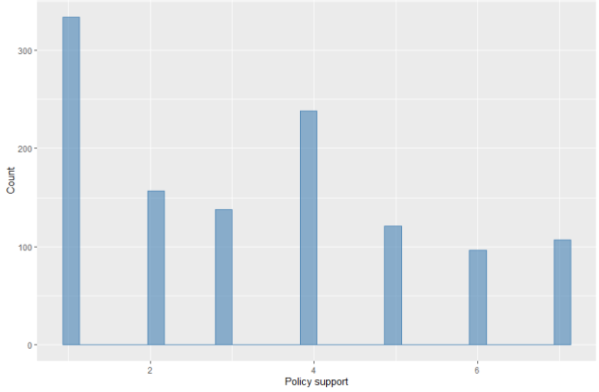

#### Potential moderators - numeracy, prosocial attitudes, and political orientation

As for Experiment 1 we explored possible interactions between quality of evidence levels and numeracy, prosociality, and political orientation on our various outcome measures.

**Numeracy:**

Numeracy in Experiment 2 was assessed using the same measures as described in Experiment 1. As in Experiment 1, we find a significant interaction between numeracy and quality of evidence level for perceived trustworthiness ($F(2,1183) = 4.64$, $p = .010$, $\eta_G^2 = 0.008$), i.e. the effect of quality of evidence level on perceived trustworthiness depends to an extent on numeracy. For lower numeracy levels the effect of quality of evidence (i.e. the difference that low quality of evidence information makes compared to high and no quality of evidence information) is less pronounced compared to higher numeracy levels. As for Experiment 1, this could hint to different levels of engagement with or understanding of the information in the infographic for people with lower numeracy compared to higher numeracy. The higher the numeracy the more pronounced the experimental effect of quality of evidence, i.e. lower trust for low quality of evidence information and higher trust for high quality of evidence information or information that does not mention quality of evidence. We do not find a significant interaction for perceived effectiveness of eye protection ($F(2,1183) = 1.50$, $p = .224$), nor behavioural uptake intent ($F(2,1183) = 0.12$, $p = .889$).

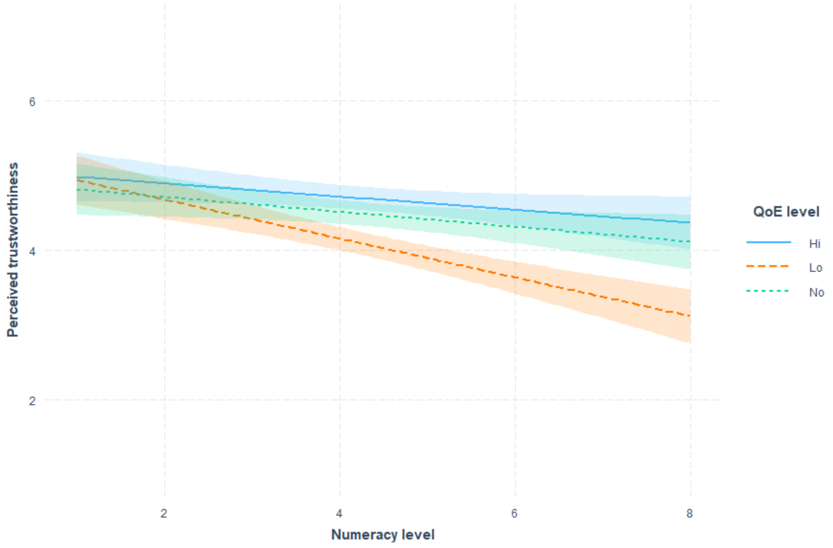

Note: Interaction plot of quality of evidence levels and numeracy on perceived trustworthiness. Light shading around trend lines denotes 95% confidence band.

**Prosociality:**

The same single item measure as in Experiment 1 was used to assess prosocial tendencies. We do not find a significant interaction between quality of evidence level and prosociality on neither perceived trustworthiness (F(2,1185) = 2.21, p = .110), nor perceived effectiveness (F(2,1185) = 1.88, p = .152), nor behavioural uptake intent (F(2,1185) = 1.40, p = .248).

**Political orientation:**

We measured political orientation with the same item as described in Experiment 1.
No significant interactions between political orientation and quality of evidence level emerged, neither for perceived trustworthiness (F(2,1183) = 0.08, p = .921), nor perceived effectiveness (F(2,1183) = 2.07, p = .126), nor behavioural uptake intent (F(2,1183) = 0.10, p = .904).
